# Integration of cell-type resolved spatial proteomics and transcriptomics reveals novel mechanisms in early ovarian cancer

**DOI:** 10.1101/2025.08.25.25333715

**Authors:** Andreas Metousis, Hilary A. Kenny, Aasa Shimizu, Lisa Schweizer, Shani Ben-Moshe, Agnes Bilecz, Rahul Krishnan, Jingwen Zhang, Isabel Alcazar, Lucy Kelliher, Mallika Ravi, Tejas Samantaray, Sabrina Richter, Yan Li, Jiying Wang, Sophia Steigerwald, Fabian J. Theis, Florian A. Rosenberger, Thierry M. Nordmann, S. Diane Yamada, Ricardo Lastra, Matthias Mann, Ernst Lengyel

**Affiliations:** Proteomics and Signal Transduction, Max Planck Institute of Biochemistry, Martinsried, Germany; Department of Obstetrics and Gynecology/Section of Gynecologic Oncology, The University of Chicago, Chicago, IL, USA; Institute of Computational Biology, Computational Health Center, Helmholtz Munich, Neuherberg, Germany; Center for Research Informatics, The University of Chicago, Chicago, IL, USA; Department of Medical Biochemistry and Biophysics, Karolinska Institutet, Solna, Sweden; Department of Pathology, The University of Chicago, Chicago, IL, USA

## Abstract

High-grade serous carcinoma (HGSC) is the most common ovarian cancer subtype, typically diagnosed at late stages with poor prognosis. Understanding early molecular events driving HGSC progression is crucial for timely detection and development of effective treatment strategies. We performed and integrated spatial cell-type resolved proteomics and paired transcriptomics across 25 women with precursor lesions of the fallopian tube and/or HGSC. Epithelial cell signatures revealed early activation of SUMOylation machinery, increased ATR and Wnt signaling, and enhanced MHC-I antigen presentation along the disease trajectory. The stroma exhibited extracellular matrix remodeling and interferon-mediated inflammation. Serous tubal intraepithelial carcinomas (STICs) in cancer patients contained a pro-coagulative signature and reduced APOA1/2 compared to STICs in individuals without cancer. We functionally established important roles of epithelial-derived TRIP13 and SUMOylation, and cancer-associated fibroblast-derived SULF1 and BGN in HGSC progression. These findings provide unique molecular insights into HGSC pathogenesis and identify potential new therapeutic targets for intervention.

## Introduction

Ovarian cancer (OvCa) is the deadliest gynecological malignancy. It is usually diagnosed at an advanced, metastatic stage, when the 5-year overall survival is only 50%^1,2^. Of the four main subtypes of OvCa, high-grade serous carcinoma (HGSC) is the most prevalent. Histologically, this tumor is characterized by diffuse nuclear atypia and near-universal p53 mutations^3–5^. While HGSC was long thought to arise from the ovarian surface epithelium^6^, this paradigm shifted with the identification of dysplastic lesions in the fallopian tube (FT) epithelium of women at increased risk for OvCa^7,8^. These lesions, characterized by aberrant p53 expression and an elevated Ki-67 proliferation index, were termed serous tubal intraepithelial carcinomas (STICs) and were considered as putative precursors of HGSC^9^ due to co-existence of STICs with up to 67% of primary HGSC^10^. Pathologists later identified two additional lesions of the FT, potentially involved in the histogenesis of HGSC, termed serous tubal intraepithelial lesions (STILs) and p53 signatures. STILs share histopathological features with STICs but have a lower Ki-67 proliferation index (<10%). In contrast, p53 signatures appear normal histologically but demonstrate a mutated pattern of p53 expression by immunohistochemistry^11^. Mutations in p53 are shared by the full range of lesions that precede HGSC and are believed to be among the earliest events in HGSC carcinogenesis^12–14^. Recent genomic studies show that these precursor lesions are molecularly heterogeneous, with distinct patterns of aneuploidy and consistent loss of chromosome 17, which correlates with proliferative activity and risk of progression^15^. Additionally, STICs exhibit secretory cell differentiation, and experimental models confirm that transformation of PAX8-positive fallopian tube secretory cells can give rise to HGSC^16,17^, providing evidence for a secretory cell origin of the disease.

The minute size of these lesions, with p53 signatures comprising as few as 12 mutated cells, represents a substantial technical challenge for molecular analysis^18^. Conventional non-cell-type resolved (‘bulk’) omics approaches cannot provide unambiguous insights into HGSC precursor lesions, because molecular signatures become diluted by surrounding cells. Therefore, cell type-resolved methods that specifically isolate the lesion cells of interest across different compartments are necessary. To overcome this challenge and preserve spatial context for molecular characterization, we turned to a spatial omics approach. We hypothesized that spatial proteomics in combination with spatial transcriptomics would provide unique insights into tissue biology. Recent advances in mass spectrometry (MS)-instrumentation have enabled unprecedented sensitivity, allowing protein identification from minimal input material while maintaining spatial resolution information^19^. We recently developed Deep Visual Proteomics (DVP), a technology that combines deep learning, cell segmentation, and classification with laser microdissection and ultra-high sensitivity MS, achieving unbiased proteomic analysis at single-cell type resolution^20^. Spatial proteomics directly measures functional molecules, although its inability to amplify proteins could cause it to miss low-abundance regulators^21^. Spatial transcriptomics, although more removed from cellular function, benefits from PCR amplification, resulting in better coverage of low-abundance molecules but with lower resolution^22^. We hypothesized that integrating these complementary approaches could provide a more comprehensive understanding of early HGSC development.

In this study, we employed DVP and Nanostring GeoMX spatial transcriptomics (Nanostring) to further elucidate the origins of HGSC by identifying molecular changes along the HGSC disease trajectory (Fig. 1A). We reasoned that the cell-type specific near single-cell resolution of these complementary technologies would enable us to precisely characterize the molecular drivers of HGSC development starting with precursor lesions in the FT. By integrating this multi-modal spatial omics data using Multi-Omics Factor Analysis (MOFA+)^23,24^, we were able to reveal novel mechanisms of HGSC pathogenesis, including early molecular alterations that precede invasive disease. The spatial omics technologies were validated by immunohistochemistry and functional studies, identifying drivers of transformation from normal tissue to invasive/metastatic HGSC.

**Figure 1.**
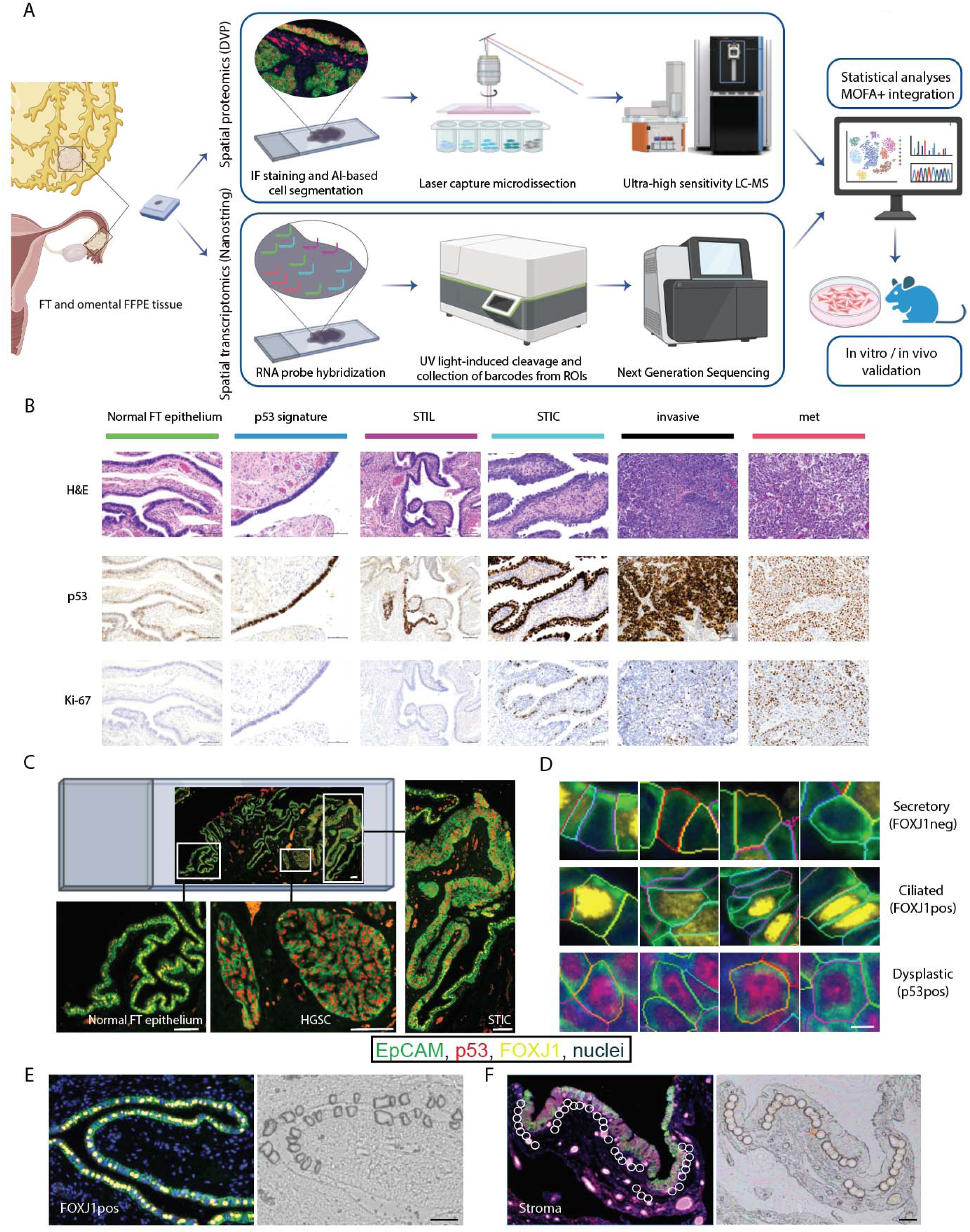
Spatial multi-omics analysis of high-grade serous cancer precursors in the fallopian tube (A) Integrated spatial multi-omics workflow. For deep visual proteomics (DVP), cell types were identified on FFPE tissue slides using antibodies and detected by immunofluorescence (IF), followed by AI-powered image analysis, laser microdissection, and ultra-sensitive MS. For Nanostring GeoMx transcriptomics (Nanostring), cell types were also identified using IF, then incubated with over 18,000 RNA probes, followed by UV-guided RNA probe cleavage and next-generation sequencing. Multi-Omics Factor Analysis (MOFA+24) integrates both datasets, followed by functional experiments. (B) Histological changes from normal fallopian tube (FT) epithelium, FT precursors, HGSC, and corresponding metastasis (met). Representative H&E staining and immunohistochemical analysis of p53 and Ki-67. Scale bar, 100 μm. (C) Representative IF images. Antibodies against EpCAM (epithelial cells, green), FOXJ1 (ciliated cells, yellow), and p53 (dysplastic cells, red) were used for cell-type identification and DVP analysis. Scale bar, 100 μm. (D) Single-cell images of secretory, ciliated, and dysplastic cells segmented by machine learning. Scale bar, 5 μm. (E-F) Representative images of FT tissue before and after laser microdissection of FOXJ1-positive (FOXJ1pos) epithelial cells (E) and STIC-adjacent stromal compartments (F). Stromal cell equivalents were detected by negative selection of cells beneath the EpCAM positive epithelial cell layers. Pre- (IF, left) and post- (bright field, right) laser capture microdissection images. Scale bar, 50 μm.

## Results

### Spatial multi-omics characterization of HGSC development from fallopian tube lesions

To investigate the molecular evolution of HGSC from its earliest precursors by multi-omics, we established a cohort of 25 women with the putative precursor lesions of HGSC, invasive FT carcinoma and/or corresponding omental metastasis (Table S1A-D). Based on consensus criteria using H&E staining and immunohistochemical analysis of p53 and Ki-67, the putative precursor lesions in the FT were classified as p53 signatures, STIL, and STIC (Fig. 1B)^11^. Invasive and metastatic HGSC were distinguished by stromal invasion and distant organ spread to the omentum, respectively.

To obtain cell type-resolved spatial proteomic and transcriptomic profiles, immunofluorescence was performed on formalin-fixed paraffin-embedded (FFPE) tissue sections. FT epithelium contains ciliated and secretory cells^25,26^, which were distinguished using ciliated lineage-specific markers FOXJ1/CAPS combined with EpCAM (epithelial cells) and p53 (dysplastic cells) (Fig. 1A, C, D, and S1A-B). For DVP, staining was followed by machine learning (ML)-powered cell segmentation and laser microdissection. In epithelial regions, we collected 150 cell shapes (15,000 µm² of 3 µm thick tissue equivalent to 50 cells) per sample (Fig. 1E), while stromal regions were captured as circular ‘cell equivalents’ totaling 25,000 µm² per sample (Fig. 1F). Bottom-up proteome profiling was performed at cell type resolution with label-free quantitation using ultra-sensitive MS. DVP analysis identified a median of 6,562 and 5,072 protein groups (distinguishable proteins, henceforth just termed proteins) per epithelial sample and stromal region, respectively (Fig. S1C). We achieved a remarkable aggregate coverage of more than 9,200 unique proteins in our dataset, with a total of over 8,500 proteins in epithelial and 6,600 in stromal samples (Table S2A, S3A). Differential protein expression analysis identified log2 fold changes up to 8 (corresponding to a 256-fold difference in abundance) between groups (Table S2B, S3B). Spatial transcriptomics was performed using the Nanostring GeoMx platform with oligo-tagged RNA probes targeting more than 18,000 genes of which about 10,000 were generally detected in samples (Fig. 1A, Table S4A). The Nanostring technology provides spatial resolution through UV-guided cleavage with a resolution down to 10 μm, capable of capturing transcripts from approximately 1-10 cell, but typically requires tens of cells for robust data^27,28^. Based on our proteomic and transcriptomic profiling, Principal Component Analysis (PCA) separated epithelial and stromal samples, validating our cell type-specific analysis approach (Fig. S1D-E).

### Spatial proteomics of the epithelium reveals stepwise molecular changes during the development of fallopian tube lesions

The median number of proteins detected was similarly high for secretory, ciliated, and dysplastic epithelial cells (Fig. S2A) with almost two thirds of the identified proteins detected in at least 75% of the samples, while high consistency across samples for each cell type and histological category was maintained (Fig. S2B-C). Normal epithelial cells, both FOXJ1-positive and FOXJ1-negative, clustered tightly, whereas dysplastic cells displayed marked heterogeneity with patient-specific clustering patterns. FOXJ1-negative secretory cells clustered closer to dysplastic cells than FOXJ1-positive cells, supporting their identification as the cell of origin for HGSC^29^ (Fig. 2A). Unsupervised hierarchical clustering analysis of epithelial samples (Fig. 2B, S2D) clearly separated STIC/invasive carcinoma cells from FOXJ1-negative/STIL/p53-signatures, driven partly by known markers of precancerous and cancerous lesions^30^ (Fig. S2E). This supports the notion that STICs are similar to invasive tumors^4^, and positions p53 signatures and STILs as very early states that retain the molecular features of normal secretory cells while harboring early transformative changes. K-means clustering yielded seven distinct protein clusters with stage-specific expression patterns (Fig. 2B, S2D, Table S2C). Overrepresentation analysis of the proteins in each cluster identified altered pathways during disease progression (Table S2D). Cluster 5, for example, includes a gradual increase in RNA processing and gene silencing proteins.

**Figure 2.**
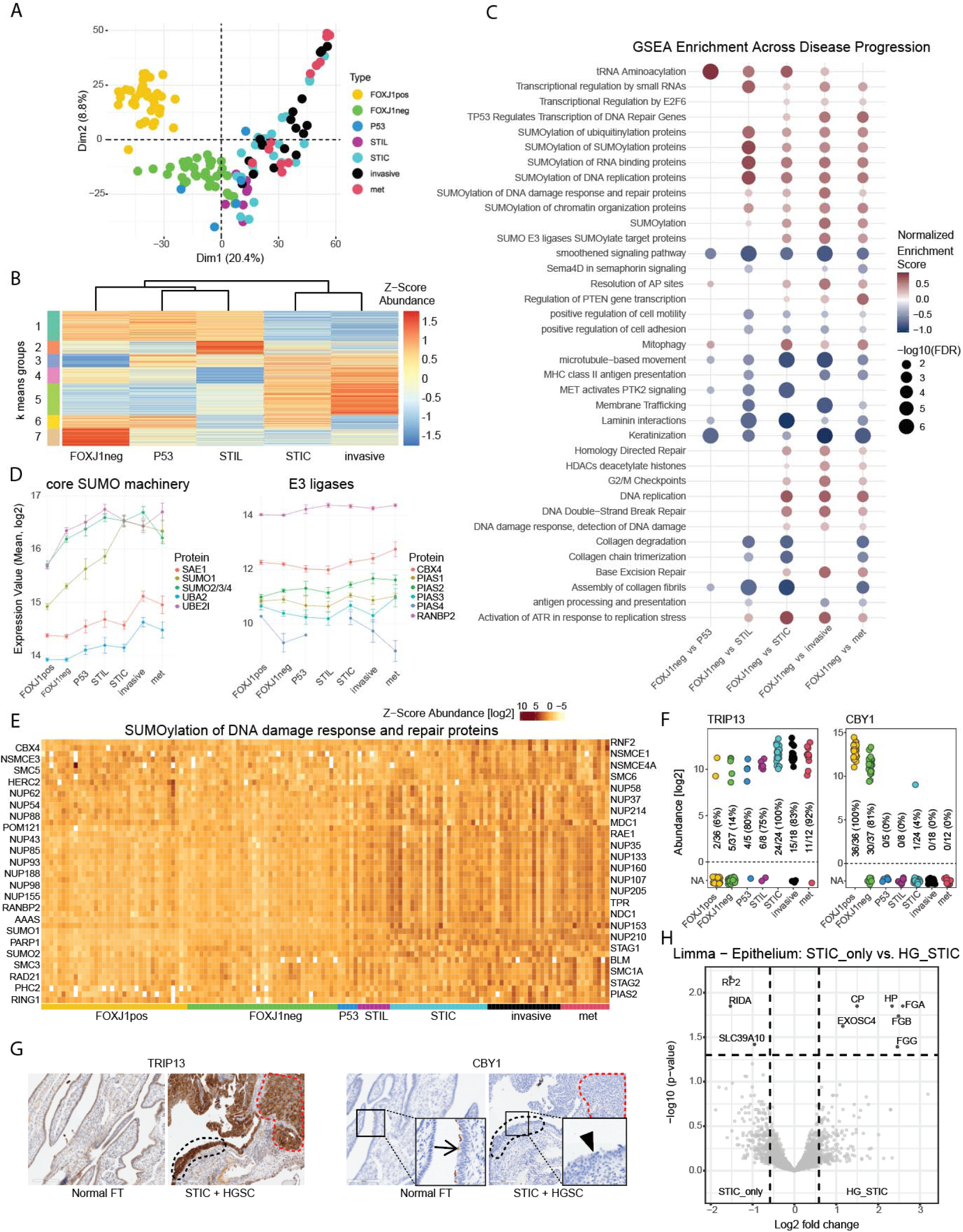
Spatial proteomics reveals progressive molecular changes during fallopian tube transformation (A) Principal component analysis of epithelial cell proteomes of normal FT epithelium, FT precursors, HGSC, and corresponding metastasis (met). FOXJ1pos, FOXJ1-positive. FOXJ1neg, FOXJ1-negative. P53, p53 signature. (B) K-means clustering analysis of epithelial proteins in seven clusters, excluding FOXJ1pos and metastasis (Table S2C). Heatmap of normalized protein abundance (z-score) across different cell states. Scale indicates relative protein expression levels. (C) Gene set enrichment analysis (GSEA) of pathway alterations during disease progression (Table S2E). Circle size indicates enrichment significance (logarithm of the FDR values); color intensity represents normalized enrichment score (fold-change). (D) Single protein abundance plots for core SUMO machinery and E3 ligases along the disease trajectory. Data shown as mean ± SEM. (E) Heatmap. Protein levels of the specific REACTOME pathway “SUMOlyation of DNA damage response and repair proteins” across the indicated sample types. Columns represent individual samples; rows represent proteins. The scale indicates normalized protein abundance. Protein names are displayed alternating between the left and right sides of the heatmap. (F-G) Protein abundance of TRIP13 and CBY1 by proteomics (F) and immunohistochemistry (G). In G: black dashed line: STIC. Red dashed line: invasive HGSC. Arrow, apical expression of CBY1 in normal FT epithelium. Arrowhead, lack of expression of CBY1 in STIC. (H) Volcano plot comparing protein expression of epithelial cells in STIC lesions from women without (STIC_only) and with (HG_STIC) concurrent HGSC (Table S2B). FDR < 0.05, fold change > 1.5.

Systematic pairwise comparisons among the marker-defined cell states (FOXJ1-positive, FOXJ1-negative, p53 signatures, STILs, STICs, invasive carcinoma, and metastatic disease) revealed thousands of differentially regulated proteins (Fig. S2F, Table S2B), serving as an important resource for studying precancerous lesions of the FT. Gene set enrichment analysis (GSEA) of these proteins uncovered pathway changes along the disease trajectory (Fig. 2C, Table S2E). Once a STIC develops, epithelial cells upregulate DNA replication and DNA damage response, including homology-directed repair and DNA double-strand break repair, which were not present in STIL or the p53-signature^31^. Interestingly, we observed increased expression of multiple SUMO pathway components starting at the STIL stage, indicating early activation of SUMOylation during FT-lesion development^32,33^. The core SUMO machinery showed coordinated and early upregulation, including SUMO proteins (SUMO1, SUMO2/3/4), E1 (SAE1, UBA2), and E2 (UBE2I) enzymes (Fig. 2D-E). Among the SUMO E3 ligases, RANBP2, which regulates proper nuclear transport and localization of DNA repair factors to damage sites, was the most strongly and earliest upregulated protein^34^. One of the early changes between FOXJ1-negative epithelial cells and p53-signature is TRIP13, an AAA ATPase^35^, which was significantly increased, while CBY1, a Wnt/Wingless pathway inhibitor^36^, was virtually absent at the proteomic level during disease progression, as confirmed using IHC (Fig. 2F-G).

When a STIC is found at a risk-reducing salpingo-oophorectomy, it is impossible to differentiate STICs that present a risk for recurrence as peritoneal carcinomatosis using a conventional H&E stain. However, women with STIC have a 28% risk of developing an invasive peritoneal carcinoma within ten years after surgery^37^. Identifying predictive markers of recurrence could be beneficial in guiding clinical decision-making. Comparing STIC lesions from women with concurrent HGSC (HG_STIC, n=7) with lesions from women without cancer (STIC_only, n=5), we identified distinct differential protein expression. Epithelium of HG_STICs showed marked upregulation of pro-coagulative and acute phase response proteins, including all three fibrinogen chains (FGA, FGB, FGG), haptoglobin (HP), and ceruloplasmin (CP), suggesting early activation of pro-inflammatory and pro-coagulative programs (Fig. 2H). In contrast, STIC_only lesions retained higher expression of RIDA, which protects cells from metabolic stress by preventing accumulation of toxic amino acid intermediates^38^ (Fig. 2H).

### Spatial proteomic analysis of the tumor microenvironment reveals stromal remodeling and inflammatory processes

Spatial proteomics of the subepithelial stroma detected a median of 5,072 proteins per sample, about 1,500 fewer than in the epithelium, likely due to the predominance of abundant proteins (e.g., collagen) which make the detection of less abundant proteins more challenging^39,40^ (Fig. S3A). Similar to the epithelial data, nearly two thirds of the identified proteins were detected in at least 75% of the samples, with high consistency across samples for each histological category (Fig. S3B-C). PCA of stromal proteomes revealed that the earliest changes occurred in STICs and became more pronounced in invasive cancers and their corresponding metastasis, while early lesions (normal-, p53 signature-, and STIL-adjacent) clustered closely (Fig. 3A). As expected, the stromal proteome of the omental metastatic site was distinct from that of all other lesions^41,42^. Unsupervised hierarchical clustering of proteins separated two main groups: Early (normal, p53 signature, STIL) *versus* late (STIC, invasive) stages (Fig. 3B, S3D), mirroring findings in the epithelial compartment (Fig. 2B), suggesting coordinated epithelial-stromal changes during disease progression. K-means clustering of these proteins resulted in seven distinct protein clusters with stage-specific expression patterns (Fig. 3B, Table S3C). Proteins from cluster 1, which were progressively upregulated, were involved in interferon signaling (IFN-α, -β, -γ) and MHC-I antigen presentation, indicating progressive immune activation during disease progression^43^ (Table S3D).

**Figure 3.**
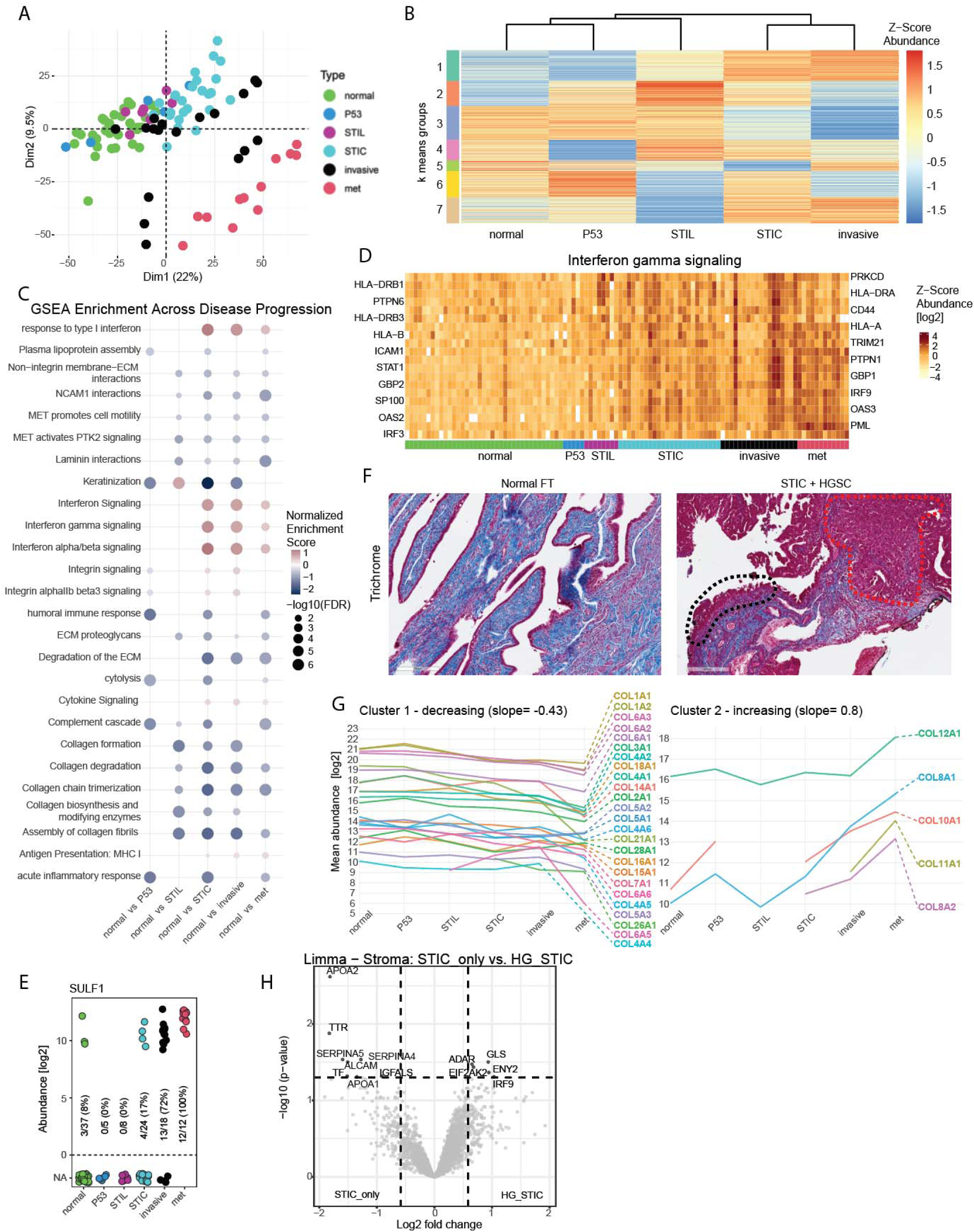
Spatial proteomics of the tumor microenvironment reveals stroma remodeling and inflammation (A) PCA of stromal proteomes under normal FT epithelium, FT precursors, and within HGSC and corresponding metastasis (met). FOXJ1pos, FOXJ1-positive. FOXJ1neg, FOXJ1-negative. P53, p53 signature. (B) K-means clustering analysis of stromal proteins in seven clusters, excluding metastasis (Table S3C). Heatmap of normalized protein abundance (z-score) across different cell states. Scale indicates relative protein expression levels. (C) GSEA of pathway alterations during disease progression (Table S3E). Circle size indicates enrichment significance; color intensity represents normalized enrichment score. (D) Heatmap showing progressive activation of interferon gamma signaling across stromal states. Columns represent individual samples; rows represent proteins involved in interferon response. Protein names are displayed alternating between the left and right sides of the heatmap. (E) Protein levels of sulfatase 1 (SULF1) across the indicated sample types determined by proteomics. (F) Representative images of collagen fibers in normal FT and in STIC that are concurrent with invasive HGSC using Trichrome staining. Black dashed line: STIC; red dashed line: invasive HGSC. (G) Decreasing (left) and increasing (right) collagen protein levels across the indicated sample types. (H) Volcano plot comparing stromal protein expression below STICs from women without (STIC_only-adjacent stroma) compared to with (HG_STIC-adjacent stroma) concurrent HGSC (Table S3B). FDR < 0.05, absolute fold change > 1.5.

Pairwise comparisons across stromal states and GSEA showed extensive protein changes during disease progression (Fig. 3C, Fig. S3E, Table S3B, E). Later disease stages exhibited increased IFN-mediated pathways, consistent with cluster 1 findings, as well as cytokine signaling (Fig. 3C-D). The extracellular matrix (ECM) began to change in STIL-adjacent stroma, showing complex remodeling of proteoglycans, laminin, neural cell adhesion molecule 1 (NCAM1), SULF1, an extracellular heparan sulfate endosulfatase, and collagens (Fig. 3C, E-G). Most collagens (COL) gradually decreased, whereas COL12A1, COL8A1, COL10A1, COL11A1, and COL8A2 increased with disease progression (Fig. 3G), consistent with roles of several of these collagens in late cancer stages and metastasis^44–47^.

Paralleling the epithelial analysis (Fig. 2H), we observed distinct stromal molecular signatures between HG_STIC and STIC_only lesions (Fig. 3H). APOA1/2 was reduced in HG_STIC stroma. SERPINA5, a coagulation inhibitor^48^, was also downregulated in HG_STIC stroma, aligning with increased coagulation factor activity seen in the epithelium. ALCAM, a cell adhesion molecule crucial for tissue organization and immune interactions^49^, was similarly downregulated, while inflammation and stress response proteins (ADAR, GLS, IRF9, EIF2AK2) were upregulated in HG_STIC stroma.

### Spatial transcriptomics provides an orthogonal layer of molecular signatures

We performed Nanostring spatial transcriptomics on serial sections from the seven epithelial and six stromal histologies previously analyzed by DVP (Fig. 1, Table S1A, C), identifying 9,742 transcripts passing quality control (QC; Fig. S4, Table S4A). While p53 signatures and STILs clustered closer to normal epithelial cells, STIC and cancer epithelium formed distinct clusters (Fig. 4A), consistent with the proteome analysis (Fig. 2A). Although ciliated (FOXJ1-positive) and secretory (FOXJ1-negative) FT cells were not transcriptionally separated by PCA (Fig. 4A), most established marker genes for these cell types^29^ were differentially expressed (Fig. S5A). The different histologic stages exhibited significant transcript changes in pairwise comparisons (Fig. S5A, Table S4B). Comparing normal secretory cells (FOXJ1-negative, hypothetical precursor cells) to STICs (Fig. 4B), alterations were observed in DNA replication, cell cycle progression, and DNA damage response. P53 stabilization also appeared among the significantly enriched pathways in STIC epithelium, along with increased Wnt signaling (Fig. 4C, Table S4C). Key Wnt-related transcripts displayed distinct patterns: WNT11, which is essential for cell migration and invasion^50^, increased at the STIC stage and remained elevated, while β-catenin (CTNNB1), a central Wnt pathway effector^51^, was already upregulated in p53 signatures (Fig. 4D).

**Figure 4.**
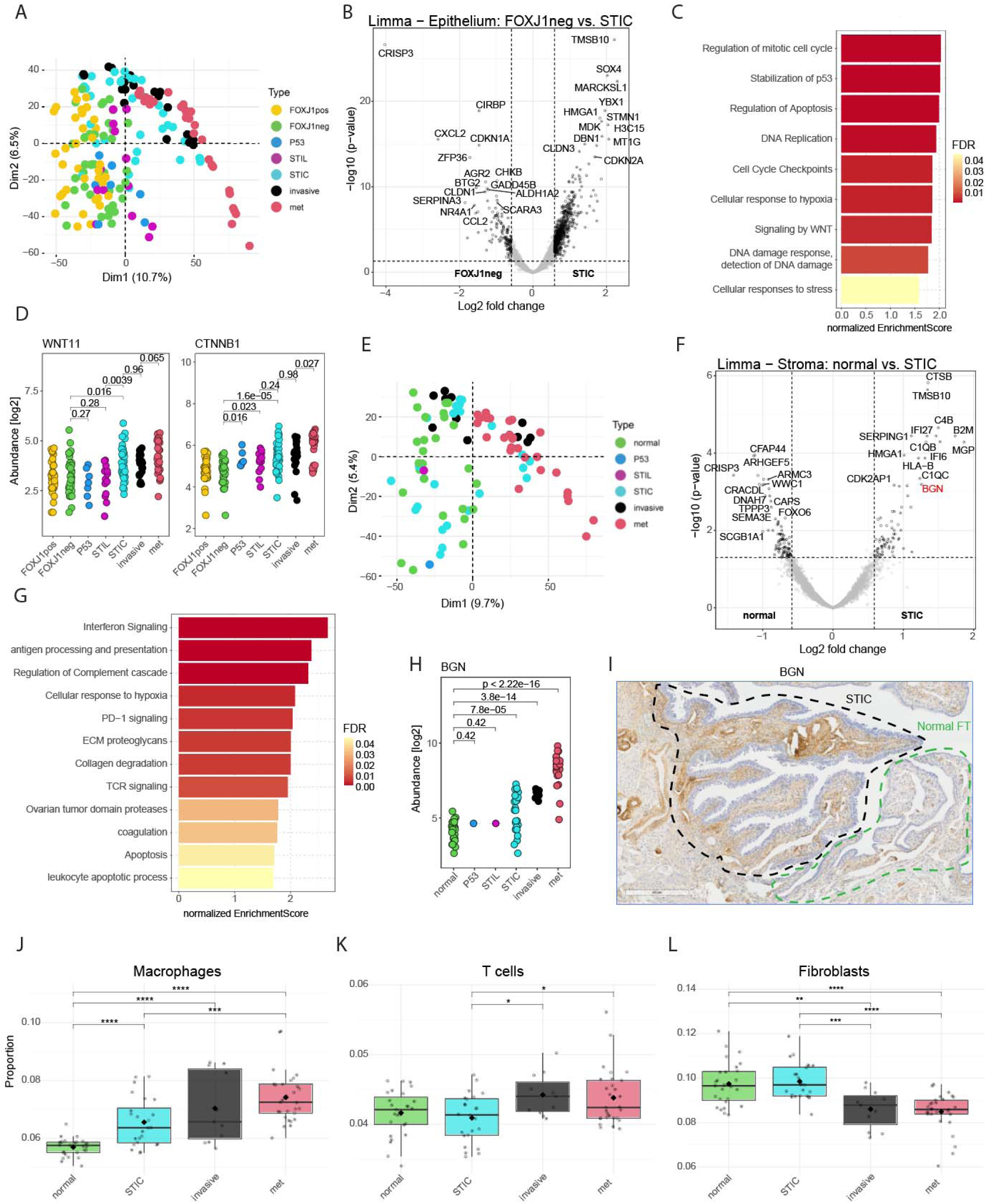
Spatial transcriptomic analysis of precancerous to HGSC transition in the fallopian tube (A) Principal component analysis (PCA) of epithelial cell transcriptomes of normal epithelium, FT precursors, HGSC, and corresponding metastasis (met). FOXJ1pos, FOXJ1-positive. FOXJ1neg, FOXJ1-negative. P53, p53 signature. (B) Volcano plot. Differential expression of transcripts between normal epithelial secretory cells (FOXJ1neg) and STICs (Table S4B). FDR < 0.05, fold change > 1.5. (C) GSEA revealing pathways of differentially expressed transcripts in STICs *versus* normal secretory cells (Table S4C). Bar length indicates enrichment significance; color intensity represents normalized enrichment score. (D) RNA expression levels of WNT11 and CTNNB1. Statistical significance was determined using unpaired two-tailed Student’s t-test. (E) PCA of stromal transcriptomes below normal FT epithelium, FT precursors, and within HGSC and corresponding metastasis. (F) Volcano plot comparing gene expression between normal and STIC-adjacent stroma, highlighting BGN and other significantly expressed transcripts (Table S4D). FDR < 0.05, absolute fold change > 1.5. (G) GSEA reveals REACTOME pathways enriched in STIC-adjacent stroma *versus* normal stroma (Table S4E). (H-I) Gene expression levels of biglycan (BGN) across the indicated sample types (H, statistics: two-tailed Student’s t-test) and immunohistochemistry in STIC-adjacent and normal stroma (black and green dashed lines, respectively) (I). (J-L) Deconvolution of Nanostring GeoMx transcriptomic data using SpatialDecon^54^ (Table S4F, G). Box plot showing the proportion of macrophages (J), T cells (K) and canonical fibroblasts (L) across disease states. Each dot represents an individual sample. Statistical significance was determined using Kruskal-Wallis test. **p < 0.01, ***p < 0.001, ****p < 0.0001. Box plots show median (center line), first and third quartiles (box limits), 1.5× interquartile range (whiskers), and mean (diamonds).

We then turned to the analysis of the stromal compartment, where the PCA showed no clear separation between STIC-adjacent and normal stroma (components 1 and 2; Fig. 4E); however, several genes were differentially expressed (Fig. 4F, Table S4D). GSEA revealed enrichment of immune-related pathways in STIC-adjacent stroma, including antigen presentation, IFN signaling, complement activation, and T-cell receptor signaling (Fig. 4G, Table S4E). Additionally, the enrichment of PD-1 signaling and leukocyte apoptotic processes in STIC-adjacent stroma suggests early development of immune evasion^52^ (Fig. 4G). The fibroblast-derived ECM proteoglycan BGN, which aligns collagen fibrils and mediates ECM signaling through direct interaction with Wnt ligands and receptors^53^, was upregulated in STIC stroma, with even stronger expression in invasive and metastatic stroma (Fig. 4H-I).

Next, we performed cell type deconvolution analysis to characterize the cellular composition of the FT stroma across disease stages (Fig. S5B, Table S4F). This revealed stage-specific alterations (Table S4G), including early upregulation of macrophages in STIC-adjacent stroma that persisted in stroma adjacent to invasive and metastatic disease (Fig. 4J). CD4+ T cells also increased in the stroma during the transition from STIC to invasive carcinoma (Fig. 4K, Fig. S5C). We observed a significant reorganization of canonical fibroblasts in invasive and metastatic disease-adjacent stroma compared to normal and STIC-adjacent stroma (Fig. 4L). These findings indicate progressive stromal remodeling with increased immune cell infiltration and fibroblast loss during FT transformation.

### Integration of DVP and Nanostring data to reconstruct disease trajectory

We employed MOFA+ to integrate spatial proteomics and transcriptomics data. MOFA+ identifies major sources of variation across different data modalities by decomposing them into shared latent factors that capture coordinated changes (Fig. 1A)^23,24,39^. This detects both shared and dataset-specific variation, providing deeper insights than single dataset analyses. We performed this analysis separately for epithelial and stromal samples to reflect their distinct biology and progression trajectories. In all epithelial samples, MOFA factor 1 captured 30% of proteomics and 10.7% of transcriptomics data variability (Fig. S6A-B). After removing FOXJ1-positive (ciliated) cells, which are not considered a HGSC precursor^16^, and metastatic cells because they represent a different anatomic region, the variability explained by MOFA1 decreased to 19.2% and 9.7%, respectively. MOFA factor 1 then clearly recapitulated the hypothesized progression from normal to dysplastic and malignant epithelial cells (Fig. 5A) as shown by the gradual decrease from FOXJ1-negative secretory cells through invasive epithelial cells (Fig. 5B). STIL and STIC were most different, implying that p53-signature and STIL are closer in protein and gene expression to normal FOXJ1 negative epithelial cells (Fig. 5B). In stroma, excluding omental metastasis, MOFA factor 1 explained 20.3% of proteomic and 6% of transcriptomic variance (Fig. 5C). It effectively separated stromal samples based on lesion progression, from normal to invasive stroma (Fig. 5C-D), again with the greatest shift between STIL and STIC. When all stromal samples, including metastasis, were included in the analysis, MOFA factor 1 did not correlate with lesion type; however, MOFA factor 2 still effectively reflected progression from normal to metastatic stroma (Fig. S6C-D).

**Figure 5.**
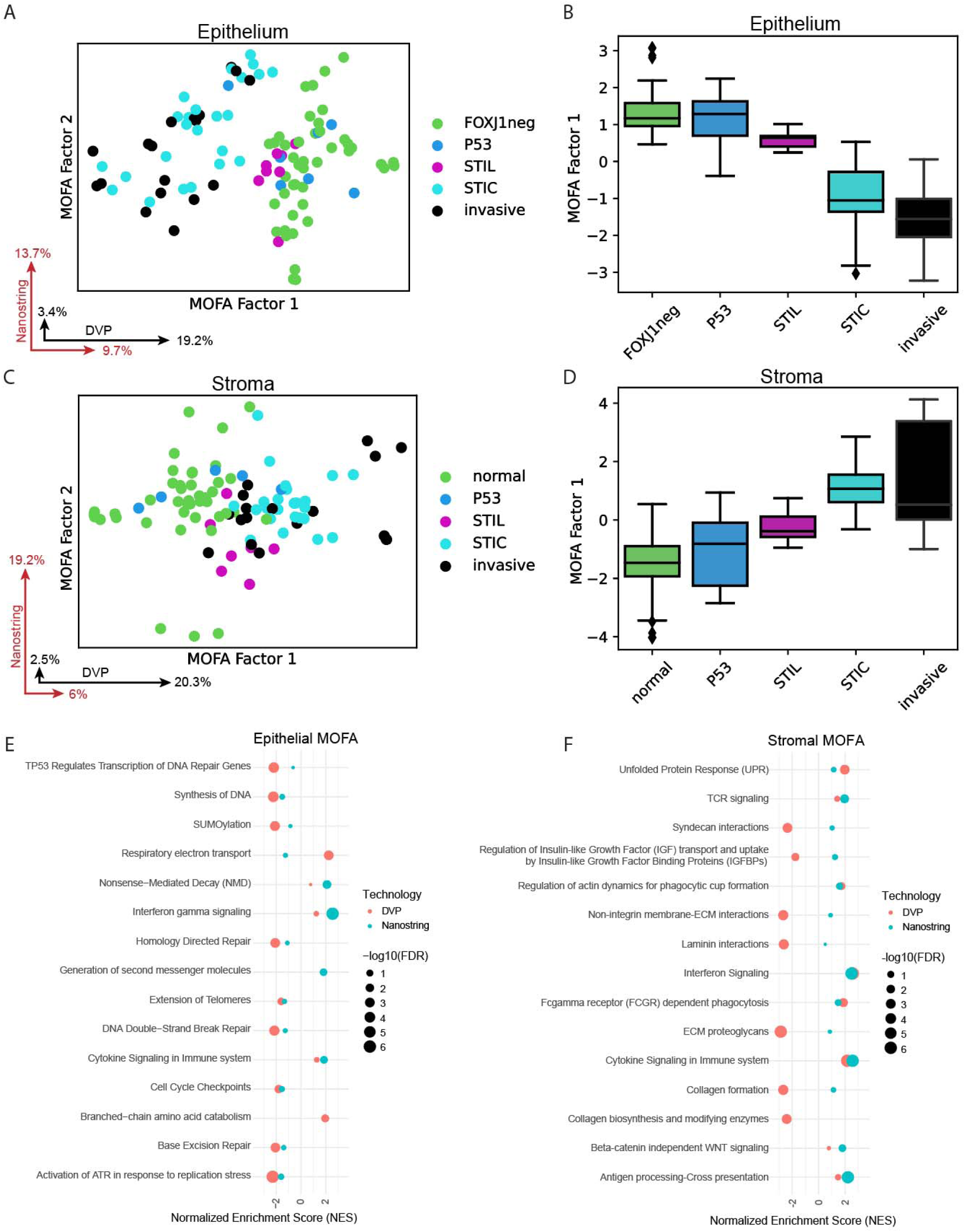
MOFA+ integration of spatial proteomics and transcriptomics reveals molecular drivers of fallopian tube lesion progression. (A-D). MOFA+ analysis^24^ excluding FOXJ1-positive (FOXJ1pos) and metastasis. The first two MOFA factors for epithelial (A) and stromal (C) samples with points representing samples; colored by cell type/lesion state. Black and red vectors show the percentage of variability of the deep visual proteomics (DVP) and Nanostring GeoMx transcriptomics (Nanostring) datasets captured by each MOFA factor. Box plots of MOFA factor 1 values across epithelial (B) and stromal (D) cell states. FT, fallopian tube. FOXJ1neg, FOXJ1-negative. P53, p53 signature. (E-F) GSEA using MOFA Factor 1 feature weights as a ranked list for proteins and transcripts in the epithelium (E) and stroma (F), excluding FOXJ1-positive (FOXJ1pos) and metastasis (Table S5I, J). Circle size is proportional to −log10 (FDR). Red, DVP. Blue, Nanostring. NES, normalized enrichment score. A positive NES indicates proteins/gene-driven pathways which are increased, and a negative NES indicates proteins/gene-driven pathways which are decreased.

We were interested in which molecular signatures contributed to these factors and analyzed feature weights for epithelial and stromal samples (Table S5A-H). To this end we generated marker panels comprising the top and bottom 10 proteins and transcripts in early and late stages, excluding or including omental metastasis because of the different tumor microenvironment (TME) (Fig. S6E-F). Notably, stromal BGN, identified in Fig. 4, ranked in the top 2.5% of transcripts, gradually increasing through the individual precancerous stages and invasive disease, while epithelial STMN1, a known STIC marker^55^, ranked first among the proteins in later stages (Fig. S6E). When metastasis was included in the analysis, BGN even ranked as the fourth highest scoring stromal transcript in advanced disease (Fig. S6F).

Using the MOFA factors from Fig. 5 A-D as input, a GSEA analysis identified significantly altered pathways linked to disease progression (Table S5I). At the epithelial level, highlights included the upregulation of pathways related to cell cycle progression, DNA replication, ATR activation in response to replication stress (Fig. 5E), and SUMOylation pathways along with DNA repair mechanisms. For stroma, increasing MOFA factor 1 values (Fig. 5D) corresponded to disease progression, with coordinated upregulation of multiple inflammatory pathways, including IFN signaling, T-cell receptor signaling, cytokine signaling, and antigen processing/cross-presentation (Fig. 5F, Table S5J).

### Therapeutic targeting of epithelial and stromal pathways and targets in ovarian cancer

Together, the above spatial analysis provided us with key pathways and targets potentially important to the cancer-related functions of OvCa cells and cancer associated fibroblasts (CAFs) (Figs. 2-5). We next wanted to investigated whether inhibiting them would have antiproliferative or other anti-tumor effects. Indeed, focusing first on epithelial targets, we found that SUMOylation inhibition (Tak-981), TRIP13 knockdown (RNAi), and TRIP13 inhibition (DCZ0415) all impaired the proliferation and invasion of OVCAR8 and OVCAR4 cells (Fig. 6A-F, S7A-E), highlighting two promising cancer cell therapeutic targets.

**Figure 6.**
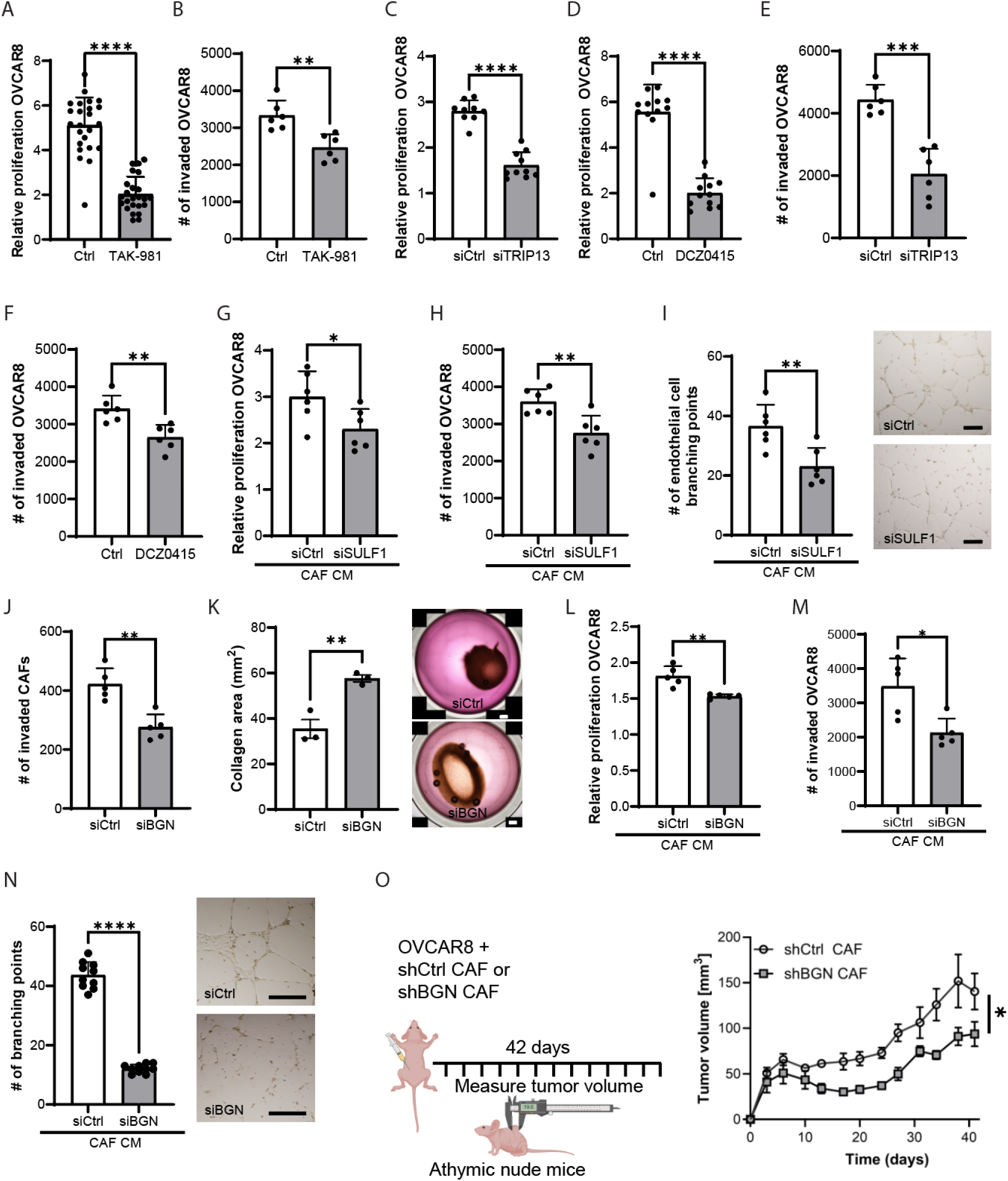
Functional validation of key molecular targets identified through spatial multi-omics (A-B) OVCAR8 cells were treated with a SUMOylation inhibitor, TAK-981, and proliferation (A) or invasion (B) was quantified. (C-F) OVCAR8 cells were transfected with control siRNA (siCtrl), TRIP13 siRNA (siTRIP; C, E) or treated with a TRIP13 inhibitor, DCZ0415 (D, F), and proliferation (C-D) or invasion (E-F) was measured. (G-I) Cancer-associated fibroblasts (CAFs) were transfected with sulfatase 1 siRNA (siSULF1) or siCtrl and conditioned media from the CAFs (CAF CM) was collected. OVCAR8 cells were treated with the CAF CM, and proliferation (G) or invasion (H) quantified. Endothelial cell branching assay including microscopy bright-field images (I). HUVEC cells were cultured on Geltrex matrix and treated with the CAF CM. Scale bar, 100 µm. (J-N) CAFs were transfected with siCtrl or biglycan siRNA (siBGN) and CAF invasion (J), CAF collagen contractility (K), or CAF CM was measured/collected. OVCAR8 cells were treated with the CAF CM from siCtrl or BGN siRNA (siBGN) transfected CAF, and proliferation (L) or invasion (M) was quantified. Endothelial cell branching assay including microscopy bright-field images. HUVEC cells were cultured on Geltrex matrix and treated with the CAF CM from siCtrl or siBGN transfected CAF. Bar graphs show Mean ± SD from three independent experiments: ** p < 0.01, *** p < 0.001, **** p < 0.0001 by unpaired t-test. Scale bar, 100 µm (endothelial cell branching assay), 1000 µm (collagen contractility assay). (O) OVCAR8 and CAFs stably expressing control shRNA (shCtrl CAF) or BGN shRNA (shBGN CAF) were co-injected subcutaneously in athymic nude mice. Tumor growth was measured over 42 days. Mean ± SD; * p<0.05 by unpaired t-test.

Second, we examined the functional role of stromal cell targets in regulating CAF activity and CAF-mediated effects within the TME. Knockdown of SULF1 in CAFs decreased the ability of CAF-conditioned media to promote proliferation and invasion of OVCAR8 and OVCAR4 OvCa cells (Fig. 6G, H, S7G, H). Conditioned media from CAFs with lower SULF1 expression was less effective at inducing endothelial cell branching (Fig. 6I). Importantly, SULF1 knockdown did not affect the proliferation or invasion of CAFs themselves (Fig. S7I, J), indicating a selective impact on the pro-tumorigenic paracrine functions of CAFs rather than their intrinsic growth or motility. We also evaluated the effects of BGN knockdown in CAFs (Fig. S7K). BGN suppression reduced CAF migration and impaired collagen contractility, key aspects of stromal remodeling, but did not alter CAF proliferation rates (Fig. 6J-K, S7L). Furthermore, BGN knockdown in CAFs disrupted CAF-conditioned media-driven proliferation and invasion of OVCAR8 and OVCAR4 cells (Fig. 6L, M, S7M, N), as well as CAF-driven endothelial cell branching (Fig. 6N). To assess the in vivo relevance of these findings, we co-injected CAFs stably expressing BGN-targeted shRNA (Fig. S7K) along with OVCAR8 cells into mice. Tumor growth was significantly decreased in mice co-injected with BGN-silenced CAFs compared to those receiving control CAFs (Fig. 6O). Therefore, BGN expression in stromal cells significantly promotes tumor growth *in vivo*.

We conclude that our integrated molecular approach combining spatial proteomics and transcriptomics identified several targets in the epithelial and stromal compartments with therapeutic relevance.

## Discussion

For this study, we developed an integrated spatial multi-omics approach, combining spatial proteomics and spatial transcriptomics, to create a comprehensive molecular atlas of HGSC progression from its earliest described putative FT precursor lesions. The technical innovations presented here address long-standing challenges in the characterization of microscopic FT precursor lesions, such as p53 signatures and STILs. Our DVP methodology overcomes previous limitations by leveraging AI-driven single-cell identification combined with machine learning, precise laser capture microdissection, ultra-sensitive MS, and multimodal data integration using MOFA+^23,24^. This enabled a deep, quantitative proteome profiling of 8,500 and 6,600 proteins in epithelial and stromal compartments, respectively, from tissue regions containing as few as 50 cell equivalents – an exceptional depth of protein analysis from a minute amount of sample^39,56^.

Our findings offer previously unattainable mechanistic insights into HGSC pathogenesis with spatial resolution. They reveal early activation of the SUMOylation machinery within the epithelium and ongoing stromal remodeling. Importantly, we identified unique molecular signatures in STICs that differentiate between progressive and non-progressive lesions. The MOFA+ integration provided complementary insights across different modalities: transcriptomics indicated Wnt signaling activation in epithelial cells, while proteomics specifically captured ECM remodeling by directly measuring extracellular proteins and more clearly highlighted early SUMOylation activation. This multimodal approach emphasizes the paired nature of these technologies and establishes a framework for studying other microscopic lesions where spatial context is essential.

Our data provides strong molecular evidence that FOXJ1-negative secretory cells are the likely cell of origin for HGSC, with these cells clustering closer to dysplastic cells than FOXJ1-positive ciliated cells. However, a recent mouse study has indicated that Krt5^+^/Prom1^+^/Trp73^+^ pre-ciliated transitional cells may be implicated in the initiation of HGSC, by showing that they are particularly susceptible to transformation upon Trp53 and Rb1 inactivation^57^. That these findings do not align with ours may be attributable to interspecies differences and varying experimental designs; notably, our human tissue analysis did not include transitional cell markers, underscoring the ongoing challenges we face in our attempts to elucidate the exact origins of HGSC. Importantly, early precursor lesions such as p53 signatures and STILs demonstrated intermediate molecular phenotypes between normal secretory FT cells and STIC lesions, yet remained more closely related to normal secretory cells. Our results suggest that p53 signature and STILs are too early in the trajectory of malignant progression to unambiguously represent precancerous lesions. Instead, our data highlight the STIL-to-STIC transition as the pivotal event in malignant transformation to HGSC, meriting more focused investigation. STICs already have many molecular features of HGSC. The widespread transcriptional and proteomic similarities among STICs, invasive, and metastatic samples further support the position of STICs as immediate precursors to invasive disease^4^.

Our analysis also uncovered distinct molecular signatures in STIC lesions from women with concurrent HGSC compared to those without cancer, providing important insights into the features of STICs with malignant potential. STIC with concurrent HGSC (HG_STICs) were characterized by a pro-coagulative and pro-inflammatory phenotype, including marked upregulation of fibrinogen chains and acute phase proteins. The reduced stromal APOA1/2 levels in HG_STICs may indicate decreased cholesterol transport to the liver and enhanced local retention for energy production. If so, this altered cholesterol metabolism^58^ would suggest that progressive STICs may be vulnerable to interventions targeting the mevalonate pathway. Indeed, statins have been shown to prevent STIC formation in mouse models^59^, raising the possibility that the altered APOA1/2 expression we observed in HG_STICs may create a therapeutic vulnerability for chemoprevention in high-risk populations. These processes are likely to contribute to the transformation of STICs, consistent with mounting evidence that both inflammation and coagulation play causative roles in carcinogenesis and therefore may have utility for risk stratification in women with incidentally discovered STICs^60,61^. RIDA expression mitigates the accumulation of reactive intermediates that damage DNA and proteins, thereby preventing the metabolic stress that could promote transformation^38^. Thus, its persistence in STIC_only lesions may confer a protective effect. The differences that we identified between HG_STIC and STIC_only lesions likely represent drivers of progression to invasive cancer.

Our results notably revealed early and coordinated activation of the SUMOylation machinery, which began at the STIL stage. The core SUMO components (SUMO1, SUMO2/3/4), E1 enzymes (SAE1, UBA2), E2 (UBE2I), were all upregulated, prominently including RANBP2. This E3 ligase is implicated in nuclear transport and localization of DNA repair factors, which may represent an adaptive initial response to accumulating DNA damage^34^. This same mechanism could later support cell survival and proliferation in conditions of genomic instability - a double-edged sword that nonetheless facilitates oncogenic transformation. This duality is shown by the simultaneous increase in cell cycle and DNA repair pathways in our MOFA+ analysis and is consistent with findings in other cancers where SUMOylation promotes cancer cell growth survival^33^. Importantly, our functional validation demonstrates that SUMOylation inhibition significantly reduces OvCa cell proliferation and invasion, consistent with SUMOylation vulnerabilities recently identified in OvCa^62^. Similarly, the progressive activation of ATR signaling in response to replication stress within the epithelium indicates that DNA damage response pathways are central to early development of HGSC. Both SUMOylation and ATR pathways likely act as parallel adaptive responses, allowing pre-malignant cells to tolerate genomic instability, a hallmark of HGSC progression^63^. This correlates with the high prevalence of homologous recombination deficiency in HGSC and supports the potential application of SUMOylation inhibitors or ATR pathway blockers-several of which are undergoing clinical evaluation [Clinical trial IDs: NCT03648372, NCT03682289, NCT02278250, NCT02223923].

The analysis of the stromal compartment revealed that microenvironmental remodeling begins early in lesion development, rather than as a consequence of invasion. Spatial proteomics allowed us to study ECM remodeling in depth during the transition from normal secretory cells to invasive HGSC. Progressive ECM changes-particularly complex collagen remodeling and the early downregulation of laminins-indicate a weakening of tissue architecture alongside epithelial transformation. While most collagens decreased, specific types (COL12A1, COL8A1, COL10A1, COL11A1, COL8A2) instead increased dramatically with progression, consistent with their known roles in advanced cancer and metastasis^44–47^. In particular, the fibroblast-derived ECM proteoglycan BGN^53^ was markedly elevated between normal and STIC-adjacent stroma. We functionally validated that BGN knockdown in CAFs reduced tumor-promoting functions and angiogenic support. Furthermore, CAFs with BGN knockdown significantly impaired tumor growth *in vivo*. BGN’s interactions with Wnt ligands and receptors are a molecular link to the Wnt pathway activation observed in epithelial cells - a relevant finding since WNT7A signaling directly drives FGF1 expression and tumor progression in OvCa^64,65^. The widespread stromal and ECM changes we observed below STIC and invasive cancer cells, compared to the stroma below normal FT epithelium, demonstrate that the FT microenvironment actively contributes to transformation. Similarly, in esophageal squamous cell carcinoma, a symbiosis of stroma and epithelial cells promotes early invasion and progression through ECM remodeling at the epithelial-stromal cell interface^66^.

Our results are supported by recent spatial transcriptomics and multiplexed immunostaining studies that have examined the early events in HGSC precursor lesions. For example, Garcia et al.^31^ provided convincing evidence that mesenchymal stem cells present in aged or BRCA-mutated FTs promote epithelial cell proliferation and survival, thereby facilitating malignant transformation. Their work highlighted oxidative stress and lipid peroxidation as key driving mechanisms of progression^31^. Building on the detailed characterization by Kader et al^43^ of immune landscape remodeling during neoplastic progression, our analysis further reveals early immune engagement within precursor lesions. Specifically, we observed activation of interferon signaling and upregulation of MHC-I at early stages, consistent with the initiation of an innate immune response. Our cell type deconvolution supported this, showing early macrophage infiltration that persists throughout disease progression, along with a significant increase in CD4+ T cells in STIC and invasive carcinoma stroma, in agreement with previous observations^43^.

Our findings have significant translational potential. Unique molecular signatures that differentiate progressive from non-progressive STICs could facilitate the development of biomarkers for risk assessment. Our functional validation of the identified targets, especially demonstrating that BGN targeting inhibits angiogenesis in vitro and reduces tumor size in vivo, provides proof-of-concept for disrupting the cancer-supporting TME. Our integrated spatial multi-omics approach delivers comprehensive molecular insights into the development of HGSC from its FT origins. Overall, our study uncovers previously unknown mechanisms of disease progression, particularly the coordinated interaction between epithelial and stromal remodeling. We also identify key molecular drivers - including SUMOylation, ATR signaling, TRIP13, BGN, and SULF1 - that drive these pathological changes. These findings clarify critical pathways involved in disease advancement and highlight promising biomarkers and potential therapeutic targets for early detection and intervention in this devastating disease.

## Limitations of the Study

Although this work represents the largest molecularly profiled collection of FT lesions with paired MS-based proteomic and transcriptomic analysis, several limitations should be considered. Because the number of cells examined from each lesion was relatively small, we may have overlooked intra-tumoral heterogeneity that could impact disease progression. While our cohort of 25 subjects is significant for this type of spatial analysis, larger studies will be necessary to fully understand inter-patient variability and identify definitive biomarkers. Additionally, our analysis did not include markers for transitional cells, which recent mouse studies suggest may play a role in the development of HGSC^57^. Future longitudinal studies with larger groups, more comprehensive marker panels, and validation in separate populations will be crucial to applying these findings clinically.

## Data Availability

Spatial proteomics data are deposited in the PRIDE database (ProteomeXchange: PXD062349) and spatial transcriptomics data in GEO (GSE305176). Pseudonymized datasets will be publicly released upon publication.

## Acknowledgements

We thank Gail Isenberg (University of Chicago) for editing the manuscript. We also thank Peter Murray (Research Group Immunoregulation, Max Planck Institute of Biochemistry) for critically reading the manuscript and providing helpful comments. Pieter Farber, Ph.D., from the U of C Genomics Facility, assisted with operating the GeoMx Digital Spatial Profiler for spatial transcriptomics.

This work was supported by the Max Planck Society for the Advancement of Science. Andreas Metousis is supported by a PhD scholarship from the Onassis Foundation (Scholarship ID: F ZS 031-1/2022-2023) and by an interdisciplinary life science fellowship awarded by the Joachim Herz Stiftung. This research was funded by an NIH/NCI R35 grant (CA264619, E Lengyel), generous philanthropic donations from Linda Usher and family (SD Yamada), Bears Care - the charitable beneficiary of the Chicago Bears Football Club (H. Kenny and E. Lengyel), and the Janet Burros Foundation (E. Lengyel).

## Author contributions

The study was conceived by E.L. and M.M. Deep Visual Proteomics (DVP) experiments were performed by A.M. with support from L.S., A.B., and I.A. Mass spectrometry measurements and optimization were performed by A.M. and S.S. The patient cohort was identified by R.L., A.B., and S.D.Y., with pathological expertise provided by A.B. and R.L. Spatial transcriptomics measurements and optimization were performed by A.S., A.B., and R.K. Omics data were analyzed by A.M., L.S., S.B.-M., F.A.R., and T.M.N. Data interpretation was performed by A.M., L.S., H.A.K. and E.L. Advanced multiomics data integration was performed by S.R. under the supervision of F.J.T. Cell type deconvolution from stromal omics data was conducted by Y.L. and J.W. Functional experiments were designed by H.A.K., A.S., and A.M., and performed by H.A.K., A.S., J.Z., L.K., M.R., and T.S. A.M. prepared the figures and wrote the original draft with E.L. and H.A.K. L.S. and MM edited the manuscript. All authors reviewed, provided feedback, and approved the final version.

## Experimental model and study participant details

### Patient cohort and ethics

Our cohort consists of 25 women who underwent surgery performed by a gynecologic oncologist in the Department of Obstetrics and Gynecology/ Section of Gynecologic Oncology at the University of Chicago and were reviewed by a board-certified gynecologic pathologist (RL, AB). For DVP, the cohort includes 24 women, five women with p53 signatures, eight with STILs, five with STIC_only, and seven with HG_STIC, four of whom also had matching omental metastases. For Nanostring, the cohort includes 13 women (12 patient-matched with DVP), three women with p53 signatures, two with STILs, two with STIC_only, and six with HG_STIC, three of whom also had matching omental metastases. All women who underwent either prophylactic salpingo-oophorectomy due to increased genetic OvCa risk or debulking surgery for potentially malignant disease gave written consent in accordance with the Declaration of Helsinki (Table S1). Samples were selected from the University of Chicago OvCa database^67^. The study has received IRB/Ethics approval from the University of Chicago and the Max Planck Institute.

## Method details

### Tissue sectioning

Archival FFPE tissue blocks were retrieved from the University of Chicago tissue biobank and sectioned at a thickness of 3µm using a microtome (Thermo Fisher Scientific, HM 340E). The slides were pretreated with UV light (254 nm) for 1 hour, immersed in 100% acetone for 5 minutes, coated with VECTABOND solution for 5 minutes (7ml VECTABOND reagent in 350 ml acetone, Biozol, #VEC-SP-1800), and washed 3x by dipping in distilled water. Sections were mounted on laser microdissection-compatible 1.0 PEN membrane slides (MicroDissect, #MDG3P40AK) and dried overnight at 37 °C.

### Immunofluorescence for DVP

Before staining, slides were heated for 40 min at 55 °C to further increase tissue adherence and immediately deparaffinized in 100% xylol twice for 2 min. Tissue was then rehydrated in a decreasing ethanol concentration series (100%, 95%, 75%, and 30%) and finally in distilled water, twice (1 min each). To retrieve the epitopes of our fixed tissue, we performed heat-induced epitope decrosslinking in a water bath set to 90 °C for 30 minutes using a buffer containing 10% glycerol in Dako Target Retrieval Solution pH 9 (Dako, S2367)^68^. Non-specific epitopes were blocked with 5% BSA in PBS for 40 min at room temperature (RT).

Primary antibodies were diluted in Dako antibody diluent buffer (Agilent Dako, S080983-2) and incubated with the tissue overnight at 4 °C. Dilutions for immunofluorescence staining: EpCAM antibody at 1:200 (CST, #14452, clone D9S3P), 1:50 anti-p53 (Agilent Dako, #M7001, clone DO-7) and 1:200 anti-FOXJ1 (Thermo Fisher Scientific, #14-9965-82, clone 2A5). The next day, slides were washed and incubated with secondary antibodies (Thermo Fisher Scientific, #A-21147, #A-2121140 and Abcam, #ab175735, 1:400 each) and diluted in PBS-1%BSA for 1 hour at 22 °C. Tissue was then counterstained with 62.5 nM Sytox Green (Invitrogen, #S7020) in H2O at RT for 7 min, and slides were mounted with water-based mounting medium (SlowFade™ Diamond Antifade Mountant, Thermo Fisher Scientific).

### Image acquisition and analysis

Immunofluorescence-labeled tissue samples were imaged using widefield microscopy (Zeiss Axioscan 7). The objective of choice was a dry 20x/0.8 that combines imaging speed with good resolution. The excitation LED intensity for all channels was set to 100%, with the exception of the UV (385 nm) channel, which was set to 5% to avoid photobleaching of the stained tissue. The exposure time for each of the fluorophores was optimized based on the intensity of each channel (typically 1-30 msec). The acquired tiles were finally stitched together into a single scan offline, on the Sytox Green channel on the ZEN Desk software, using the default settings.

Image analysis – cell segmentation and classification. Stitched .czi files were imported to the Biology Image Analysis Software (BIAS, Single Cell Technologies), and the tiles, including epithelial and stromal components, were exported as .tiff files to be used as input for CELLPOSE. The default cyto2 CELLPOSE model, which used EpCAM as the main segmentation channel and Sytox Green as the secondary (nuclear) channel, was further improved by employing a “human in the loop” approach^69,70^ and applied to all the samples. The output masks were imported back to BIAS for further analysis. First, the duplicates were removed (overlap threshold = 25%) and then cells were classified as ciliated or secretory based on the presence (ciliated cells) or absence (secretory cells) of FOXJ1 staining. Dysplastic or normal appearing cells were classified based on the presence/absence of a bright p53 signal. The resulting epithelial cell contours were overlaid with expert tissue annotations designating normal FT areas and lesions; including p53 signatures, STILs, STICs, invasive tumors, and metastases, and assigned to their respective groups. In addition to epithelial cells, the stroma adjacent to the epithelial cells of each group was manually selected for dissection by a gynecologic pathologist (AB). The resulting contours were exported in .xml files, along with three calibration points that were placed on characteristic positions on each slide. At the same time, the optimal cutting path was defined by the “greedy” algorithm of BIAS.

### Laser capture microdissection

A total volume of 45,000 μm^3^ (3 μm thick tissue x 15,000 μm^2^ area collected) of cells were laser microdissected using a LMD7 (Leica Microsystems) and pooled per sample and group of cells, corresponding to approximately 100-150 cell shapes (or around 50 cell volumes). Adjacent to these epithelial cells, another 75,000 μm^3^ (3 μm x 25,000 μm^2^) of stroma was captured as round ‘cell equivalents’ using the LMD7. The settings used were Power = 40, Aperture = 1, Speed = 15, Middlepulse count = 1, Final pulse = −2, Head current = 51%, Pulse frequency = 2900 and Offset = 190. Samples were collected dry in LoBind 384-well plates (Eppendorf) and stored at room temperature until further processing.

### Sample preparation and mass spectrometry

Preparation of the microdissected samples for MS in the 384-well format was performed in an automated fashion utilizing a robotic liquid handling system (Agilent Bravo), as described in detail by Schweizer et al^39^. The resulting digested samples were loaded on evotips (Evosep), according to the manufacturer’s instructions.

The samples were chromatographically separated using an Evosep One running the Whisper 40 samples per day (31-minute-long) chromatographic gradient on an Aurora Elite XT column (IonOpticks, 15cm x 75μm, C18, 1.7μm particle size, # AUR3-15075C18-XT) at 55 °C and 1900V and subsequently measured on a Thermo Orbitrap Astral MS coupled with the FAIMS Pro Interface operating at a single compensation voltage of −40V. The MS instrument settings for the epithelial samples had a 240K MS1 resolution, 8 Thomson window width from 380 to 980 m/z, and 18 msec maximum injection time. For the stromal samples, the MS1 resolution was kept at 240K with a variable window approach targeting deepest proteomic coverage. Specifically, we used 60 windows spanning the range from 380 to 980 m/z and a maximum injection time of 20msec.

### Spatial proteomics data processing

Raw mass spec data (.raw files) were searched separately for epithelial and stromal samples, using DIA-NN v1.8.1^71^ on the library free mode, enabling both “FASTA digest for library-free search / library generation” and “Deep learning-based spectra, RTs and IMs prediction”. The FASTA file used was UP000005640_9606.fasta (retrieved from Uniprot, July 2021). Specifically, the rest of the settings used were as follows: up to 1 missed cleavage, up to 1 variable modification (N-term M excision, C carbamidomethylation and Ox(M)), 7-55 peptide length range, 2-4 precursor charge range, 300-1200 precursor m/z range, 200-2000 fragment ion m/z range, precursor FDR 1%, genes for protein inference, single-pass mode for neural network classifier, robust LC (high precision) for quantification strategy and global for cross-run normalization. The “MBR” and the “heuristic protein inference” boxes were also checked. From the resulting files, the “pg_matrix.tsv” was used for downstream analysis within the R statistical framework, as described in the “data analysis” section.

### Spatial transcriptomics

We performed histological and immunofluorescence (IF) analyses to characterize the specific lesions precisely. Representative hematoxylin and eosin (H&E) stained sections alongside IF images are presented in Figure 1A. For an accurate delineation of epithelial regions, we implemented a multicolor IF staining approach. Epithelial areas were identified using pan-cytokeratin (PanCK) visualized with green fluorescence, while ciliated cells were highlighted by Calcyphosin (CAPS) with yellow fluorescence. Mutated p53 expression, an established marker of early oncogenic transformation in HGSC, was detected using red fluorescence, and nuclei were counterstained with blue fluorescence. Stromal compartments were identified through negative selection. Tissue processing began with sectioning FFPE samples to a 5 μm thickness following the Nanostring GeoMx user manual (MAN-10132-04). After deparaffinization and rehydration, protein target retrieval was performed using Tris-EDTA antigen retrieval buffer for 20 minutes in a pressure cooker. RNA targets were retrieved through proteinase K digestion for 15 minutes at 37 °C. Tissues were then post-fixed in 10% NBF and hybridized overnight at 37 °C with RNA probes (Human Whole Transcriptome Atlas, containing 18,000 protein-coding genes coupled to UV-cleavable oligonucleotide barcodes). Following hybridization, excess probes were removed by stringent washing at 37 °C and incubated in blocking buffer W (Nanostring LOT#:2-23020032).

The tissue sections were subsequently stained with conjugated primary antibodies targeting pan-cytokeratin (mouse monoclonal antibody, Novus NBP2-33200AF488) and p53 (Dako primary and secondary antibodies), along with CAPS (primary and secondary antibodies) and SYTO13 nuclear stain (ThermoFisher Scientific, S7575) for one hour at room temperature.

For transcriptomic data acquisition, slides were analyzed on the GeoMx Digital Spatial Profiler (DSP) at 20× magnification. Regions of interest (ROIs) were identified based on immunofluorescence images with a pathologist’s (AB) supervision and matched to regions selected in DVP. Following ROI selection, RNA probes were photocleaved using UV illumination within each ROI, and oligonucleotide barcodes were collected into a 96-well plate, allowing precise separation of tissue compartments. Sequencing libraries were prepared using Illumina TruSeq technology for adaptor ligation and amplification. The amplified libraries underwent two rounds of Ampure XP magnetic bead cleanup (1.2:1 beads ratio) (HighPrep PCR from MAGBIO, Cat#: AC-60500). Purified libraries were sequenced on an Illumina NovaSeq flowcell PE50 at a depth of 100× per μm².

### Data analysis

Bioinformatics analyses were performed in R (version 4.3.2) on log2-transformed protein intensities from the Deep Visual Proteomics dataset and count data from Nanostring GeoMx. Data completeness was evaluated by calculating the percentage of non-missing values across all samples for each protein, categorizing them into different completeness tiers (>75%, 50-75%, and <50% completeness). For proteomics quality control, the inter-specimen coefficient of variation (CV) was assessed by calculating the standard deviation divided by the mean of intensities for each protein across different samples within the same cohort (FOXJ1pos, FOXJ1neg, P53, STIL, STIC, invasive, and metastasis). This approach quantified biological variability between specimens of the same cohort type. CV analysis was performed on a filtered subset of the dataset containing only proteins with at least 70% valid values, with distributions visualized as violin plots to compare variability patterns across cohort groups.

GeoMx raw count data was processed and normalized using the R GeoMxTools package (v3.6.2). Quality control was implemented by filtering segments based on several parameters: segments with fewer than 1000 raw reads, below ∼50% for % Aligned, ∼80% for % Trimmed, and below 50% for % Stitched sequencing reads were removed. Additionally, segments with sequencing saturation <50%, negative count <1, and No Template Control count >9000 were excluded. After applying these quality thresholds, segments with less than 2.5% of genes detected were removed. The remaining data were normalized using Q3 normalization to account for technical variation between samples while preserving biological differences.

Differential protein/transcript abundances were determined using the ‘Limma’ package (v3.58.1) with false discovery rate (FDR) for multiple testing correction and defined significance parameters (adjusted p-value ≤ 0.05 and logarithmic fold-change > 1.5). For all Principal Component Analyses (PCA), data were filtered to include proteins with at least 70% valid values in each sample group, and missing values were imputed using a sample-from-normal distribution approach (width of 0.3, downshift of 1.8). PCA was performed using the FactoMineR package (v2.11). Biological pathway enrichments were conducted through gene set enrichment analysis (GSEA) on significantly differentially regulated proteins/transcripts using the WebGestalt toolkit with reference to ‘Reactome Pathway’ and ‘non-redundant gene ontology Biological Process’ databases, applying Benjamini-Hochberg FDR correction (cutoff: 0.05). The markers defining ciliated and secretory cells were derived from the 100 most highly expressed genes in post-menopausal FT samples, based on recently published data^29^. Visualizations were generated using the ggplot2 package (v3.5.1), with p-values calculated using paired Student’s t-tests assuming equal variances.

To analyze expression trends of collagens across disease progression, we identified two collagen clusters with similar expression patterns across the different categories of stroma (“normal”, “P53”, “STIL”, “STIC”, “invasive”, and “met”). We implemented a first-to-last point slope method, which calculated the overall trend based on the difference between the expression at the first and last categories. Hierarchical clustering was performed on the calculated slopes using Euclidean distance and Ward’s method (ward.D2, cluster package, v2.1.4) to group proteins with similar expression trajectories.

K-means clustering was applied to both epithelial and stromal samples after categorizing them by biopsy type. For epithelial samples, categories included ‘FOXJ1neg’, ‘P53’, ‘STIL’, ‘STIC’, ‘invasive’, and ‘metastatic’, while stromal samples were grouped as ‘normal’, ‘P53’, ‘STIL’, ‘STIC’, ‘invasive’, and ‘metastatic’. Mean expression was calculated for each group after excluding two outlier stromal samples. Protein expression values were standardized using Z-scores, and K-means clustering was performed (K = 7, 100 random initializations) on the Z-scored matrices. Resulting clusters were visualized using the ‘pheatmap’ package. Over-representation analysis (ORA) was conducted for each protein cluster using WebGestaltR, comparing protein clusters against human Reactome and Hallmark geneset databases. Enrichment ratios, hypergeometric test p-values, and FDR-adjusted values were calculated for pathway associations within each cluster.

### Cell Type Deconvolution Analysis

To estimate the relative abundance of cell types within spatial regions, we applied SpatialDecon^54^ to the Nanostring spatial transcriptomics dataset. The raw expression matrix was normalized and preprocessed following the recommended SpatialDecon workflow. Cell type deconvolution was performed using a reference-based approach, leveraging a curated expression signature matrix derived from a published single-cell atlas of the human FT, which includes post-menopausal samples from the fimbriae region of healthy individuals^25,29^. The resulting cell type proportion estimates were compared across sample types and visualized to explore spatial heterogeneity. Due to limited sample sizes for p53 signature (n=1) and STIL (n=1), these samples were grouped with normal and STIC samples, respectively, for statistical analyses.

### Data integration – Multi-Omics Factor Analysis

Integration of transcriptomic and proteomic datasets was performed using MOFA+ framework^23,24^. MOFA+ is particularly well-suited for this study because it can handle the inherent differences in data structure, scale, and dimensionality between proteomics and transcriptomics datasets while still identifying biologically meaningful patterns across both modalities. This statistical framework identifies major sources of variation across multiple data modalities by decomposing them into a set of shared latent factors that capture coordinated changes across datasets. Unlike traditional integration methods, such as canonical correlation analysis (CCA) or partial least squares (PLS), that focus primarily on correlative relationships^72,73^, MOFA+ can detect both shared and dataset-specific sources of variation, making it ideal for our goal of both understanding common disease progression signatures and gaining modality-specific insights.

The analysis included only differentially expressed genes and proteins identified from previous analyses. For epithelial tissue, we integrated 182 Nanostring (transcriptomics) and 140 DVP (proteomics) samples, and for stromal tissue, 93 Nanostring and 104 DVP samples. When multiple measurements existed for individual samples, values were averaged before integration. The multimodal analysis was conducted separately for epithelial and stromal tissues using the Muon Python framework, a specialized tool for multimodal omics data integration^74^. This approach enables us to disentangle variation unique to specific data modalities from variation shared across modalities, providing more comprehensive insights than analyses of individual datasets alone. Four latent factors were extracted for each tissue type from the combined Nanostring and DVP datasets to account for their distinct biological properties and progression trajectories.

### Cell Culture

OVCAR8 (ATCC) and CAF immortalized cell clones^75^ were cultured in Dulbecco’s Modification of Eagle’s Medium 1X (Corning) supplemented with 1% non-essential amino acids (Corning), 1% MEM vitamins (Corning), 10% fetal bovine serum (FBS) (complete DMEM), and 1% penicillin-streptomycin. OVCAR4 (T. Hamilton, NCI-Fredrick Cancer DCTD Tumor/Cell Line Repository) cells were cultured in RPMI 1640, 1X with L-glutamine (Corning) and 10% FBS. All cells were maintained at 37 °C in a humidified incubator at 5% CO2. Cell lines were banked in liquid nitrogen and passaged at least twice after thawing before beginning experiments. Each vial was confirmed as *Mycoplasma-* negative using the STAT-Myco kit. Previously established cell lines were validated using short tandem repeat DNA fingerprinting with the AMPFℓSTR Identifier kit and compared to known fingerprints through IDEXX BioAnalytics Laboratories (Columbia, MO). Cells were passaged 2–10 times after thawing before commencing with experiments.

### Primary Cell Isolation

*Cancer-associated fibroblast isolation*. Written informed consent was given by two patients undergoing primary OvCa debulking surgery, who were diagnosed with metastatic HGSC, in compliance with University of Chicago Institutional Review Board-approved protocols. CAFs were isolated from omental tumor as previously described^76^.

### Immunohistochemistry/Trichrome stain

FFPE tissue was sectioned (5 μm) on SuperfrostTM Plus Microscope Slides (Fisher Scientific, 22-037-246). Then, the immunohistochemistry was performed on Leica Bond RX automated stainer. After the standard procedures for deparaffinization and rehydration, tissue sections were treated with Proteinase K (Agilent, S3004) for 5 minutes pre-treatment at room temperature. Anti-Ki67 (ThermoFisher, 101AP, 1:200), p53 (Millipore Sigma, pantropic, AB-12, OP140, 1:100), TRIP13 (Proteintech, 19602-1-AP, 1:300), CBY1 (Proteintech, 12239-1-AP, 1:400) and BGN (Proteintech, 16409-1-AP, 1:400), were applied on tissue sections for 60 minutes incubation at room temperature. The antigen-antibody binding was detected by Bond Polymer Refine Detection (DS9800, Leica Microsystem). Tissue sections were briefly immersed in hematoxylin for counterstaining and were covered with cover glasses. These slides were imaged using Olympus VS200 Slideview. Images for publication were exported from QuPath version 0.3.2. Trichrome staining to detect collagen fibers was conducted as previously published.^76^

### RNAi

To achieve a transient knock-down of target genes, CAFs, OVCAR4, and OVCAR8 cells were transfected with small interfering RNA SMARTPools for *BGN* (Horizon Discovery, L-021493, 25 nM), *TRIP13* (Horizon Discovery, L-016262, 25 nM), *CBY1* (Horizon Discovery, L-017047, 20 nM), or *SULF1* (Horizon Discovery, L-006643, 10 nM) using DharmaFECT 1 (Horizon Discovery). The control siRNA was siGLO RISC-Free Control reagent (Horizon Discovery; D-001600-01-05).

To achieve stable knock-down of *BGN*, CAFs were transfected with shRNA lentiviral particles (Sigma Aldrich, TRCN0000153232, TRCN0000153413). The control shRNA lentiviral particles (Sigma Aldrich, SHC002) in DMEM 1X, without FBS and penicillin-streptomycin. The media was exchanged for DMEM 1X with 10% FBS and 1% penicillin-streptomycin or other condition media 24 hours after transfection.

### Inhibitor testing

The TRIP13 inhibitor (DCZ0415^77^, OVCAR4 5 M, OVCAR8 10 M), and SUMOylation inhibitor, (TAK981^78^, OVCAR4 200 nM, OVCAR8 100 nM) were purchased from MedChemExpress. The inhibitors were tested in vitro using the proliferation IC50 doses.

### Immunoblots

Cells were plated to a 6-well plate at a concentration of 1-2 × 10^5^ cells per well and cultured for 48-72 hours before isolating protein. At this point, the cells were lysed with an SDS lysis buffer containing 4% SDS and 10 mM HEPES at a pH of 8.5, and 15-25 µg of each cell lysate was separated via SDS-PAGE on a 5-20% gel (Bio-Rad). Next, the gel was transferred to a nitrocellulose membrane (ThermoFisher) using the Power Blotter System (ThermoFisher). The membrane was then blocked with 5% non-fat dry milk (Lab Scientific) in Tris-buffered saline with 0.1% Tween 20 (American Bio) (TBST) at room temperature for 1 hour. Membranes were incubated in primary antibodies for SULF1 (ThermoFisher, PA5-115984, 1:1000), TRIP13 (Proteintech, 19602-1-AP, 1:1000), CBY1 (Proteintech, 12239-1-AP, 1:1500), BGN (Proteintech, 16409-1-AP, 1:1000), and GAPDH (Cell Signaling, 5174, 1:2000) in 5% bovine serum albumin in TBST at 4 °C overnight. After removing the primary antibody and washing with TBST, the membrane was incubated in Rabbit secondary antibody (1:2000) (Cell Signaling) conjugated to horseradish peroxidase in 5% non-fat dry milk in TBST for 1 hour at room temperature. Membranes were developed using Clarity Western ECL Substrate (Bio-Rad) and proteins were visualized with the ChemiDoc XRS+ System (Bio-Rad).

### Collection of conditioned media

Media used for functional assays was collected from CAFs. These cells were transfected with siRNA and after 24 hours the media was exchanged for serum-free media. The cells were then left to culture in this media for another 24-48 hours before collecting the media. The media was aspirated from each well and spun down so that any cellular debris was pelleted. The supernatant was then filtered through a 0.22 µm filter and either stored at −80 °C for later use or added directly to epithelial cells for a functional assay.

### Proliferation Assay

OVCAR4, OVCAR8, and CAFs transfected with or without siRNA were plated in 96-well plates at a concentration of 2,000 cells per well and left to culture for 24 hours before changing media. Next, control condition media (DMEM 1x, 10% fetal-bovine serum, 1% pen-strep), and media collected from CAFs transfected with siRNA or shRNA were added to the respective wells. The plate was then placed in the IncuCyte® Live Cell Analysis System (Sartorius) and the wells were imaged every 4 hours for 72 hours to evaluate proliferation^79^.

### Invasion Assay

OvCa cells or CAFs transfected with or without siRNA were plated in the top well of the QCMTM 96-Well Cell Invasion Assay plate (40,000 cancer cells and 4,000 CAFs/well). For siRNA testing, the cells were pre-transfected with target or control siRNA 24 hours prior to assay in serum-free media. For inhibitor testing, the cells were treated with IC50 concentration of inhibitors in serum-free media. Growth media was placed in the bottom chamber for the siRNA and inhibitor testing. The cells were incubated for 24 hours to allow the cells to invade through the chamber. The invaded cells were detached, lysed, and stained with CyQuant GR Dye according to manufacturer instructions. Total fluorescence (480/520 nm) was acquired on a fluorescent plate reader (SpectraMax iD5).

### Collagen contractility assay

Rat-tail collagen type I (0.6 mg; Corning 354236) was diluted in growth media, neutralized with 0.2N NaOH, mixed with CAFs (0.2 x 106), and plated into a 24-well dish. The CAFs were transfected with siRNA 24h prior to assay setup. The cells were incubated at 37 C° 5% CO2 for 24 hours. The collagen gel contraction was imaged and measured using a Nikon Eclipse Ti2^80^.

### Endothelial cell branching assay

Twenty-four well plates were coated with 100 μL/well of Geltrex Matrix. A total of 50,000 human umbilical vein endothelial cells were plated/well on top of the matrix-coated wells in LVES-supplemented Medium 200. After 2 hours, the media was removed, and the cells were treated with CAF316 cell CM (described above) or control growth media. After 12 hours, cell images were captured using a Nikon Eclipse Ti2. The number of branchpoints was quantified using an angiogenesis analyzer plugin of ImageJ Software.

### In vivo xenograft model

All procedures involving animal care were approved by the Committee on Animal Care at the University of Chicago. Mice were injected subcutaneously (s.c.) with 1 million OVAR8 and 6 million CAFs infected with shCtrl or shBGN cells. The mice were sacrificed 42 days post cancer cell injection. The tumors were collected, fixed in 10% buffered formalin and paraffin embedded^81^.

### Use of LLMs and generative AI

Claude (Anthropic) was used to assist with grammar checking, improving clarity of scientific writing, and organizing literature references. The authors take full responsibility for all scientific content and conclusions.

