## supplemental figures and table legends for "Integration of cell-type resolved spatial proteomics and transcriptomics reveals novel mechanisms in early ovarian cancer"

Supplementary figures
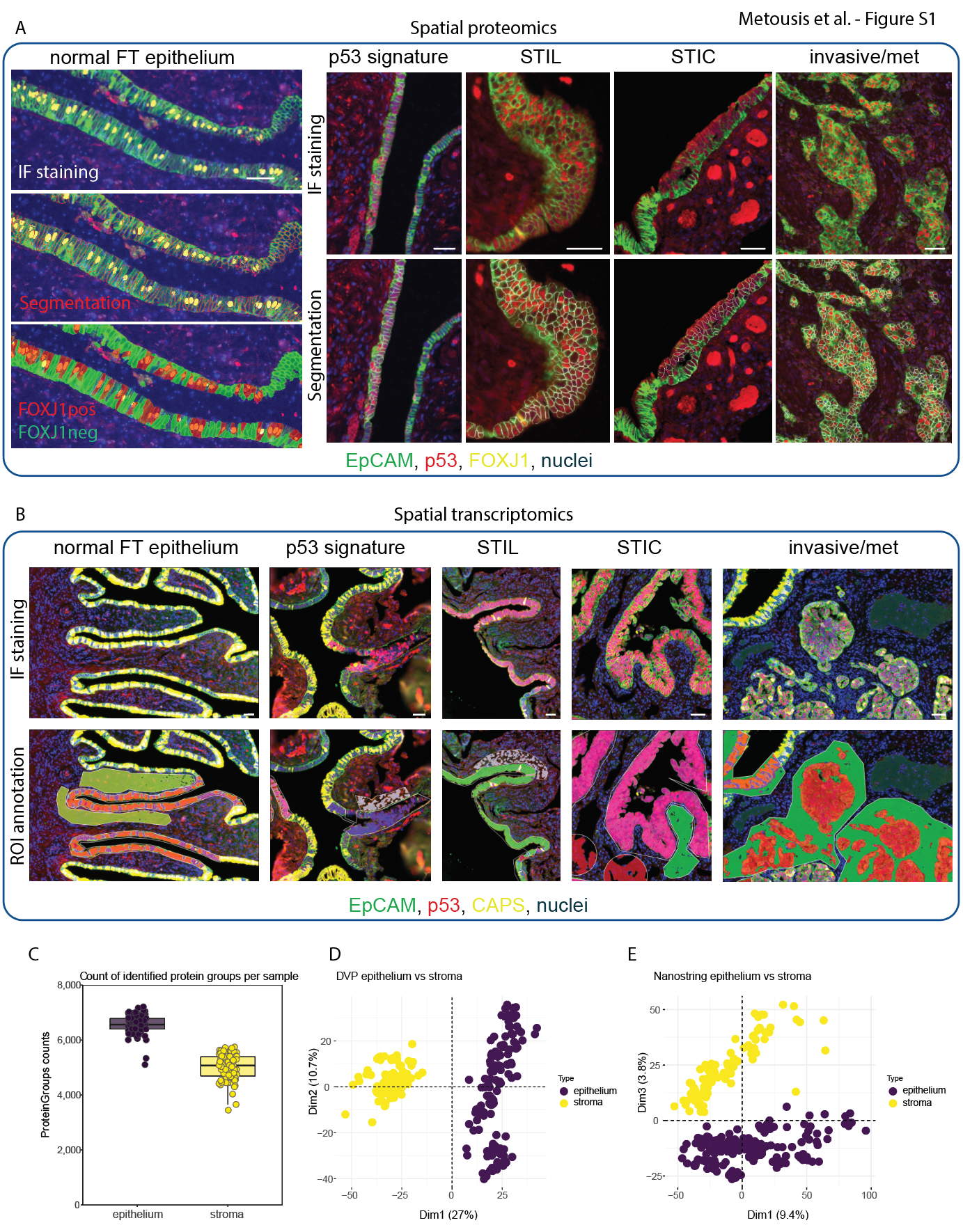
 **Figure S1. Complementary spatial proteomics and transcriptomics characterize fallopian tube lesion progression, related to Fig. 1**

(A) Deep visual proteomics (DVP). Cell types are identified on FFPE tissue slides by immunofluorescence (IF), followed by AI-powered image analysis for cell segmentation (white outlines). Scale bars, 50 μm. FOXJ1pos, FOXJ1-positive cells. FOXJ1neg, FOXJ1-negative cells. met, metastasis.

(B) Nanostring GeoMx transcriptomics (Nanostring). Cell types are identified on FFPE tissue slides by IF, and region of interest (ROI) annotations for targeted probe collection are selected. Scale bars, 50 μm.

(C) Median count of identified protein groups across epithelial (purple) and stromal (yellow) regions adjacent to each lesion type. Box plots show median, quartiles, and range; individual data points represent single measurements.

(D-E) Principal component analysis demonstrates clear separation between epithelial and stromal samples in DVP proteomics (D) and Nanostring transcriptomics (E) datasets, validating the cell type-specific analysis approach.

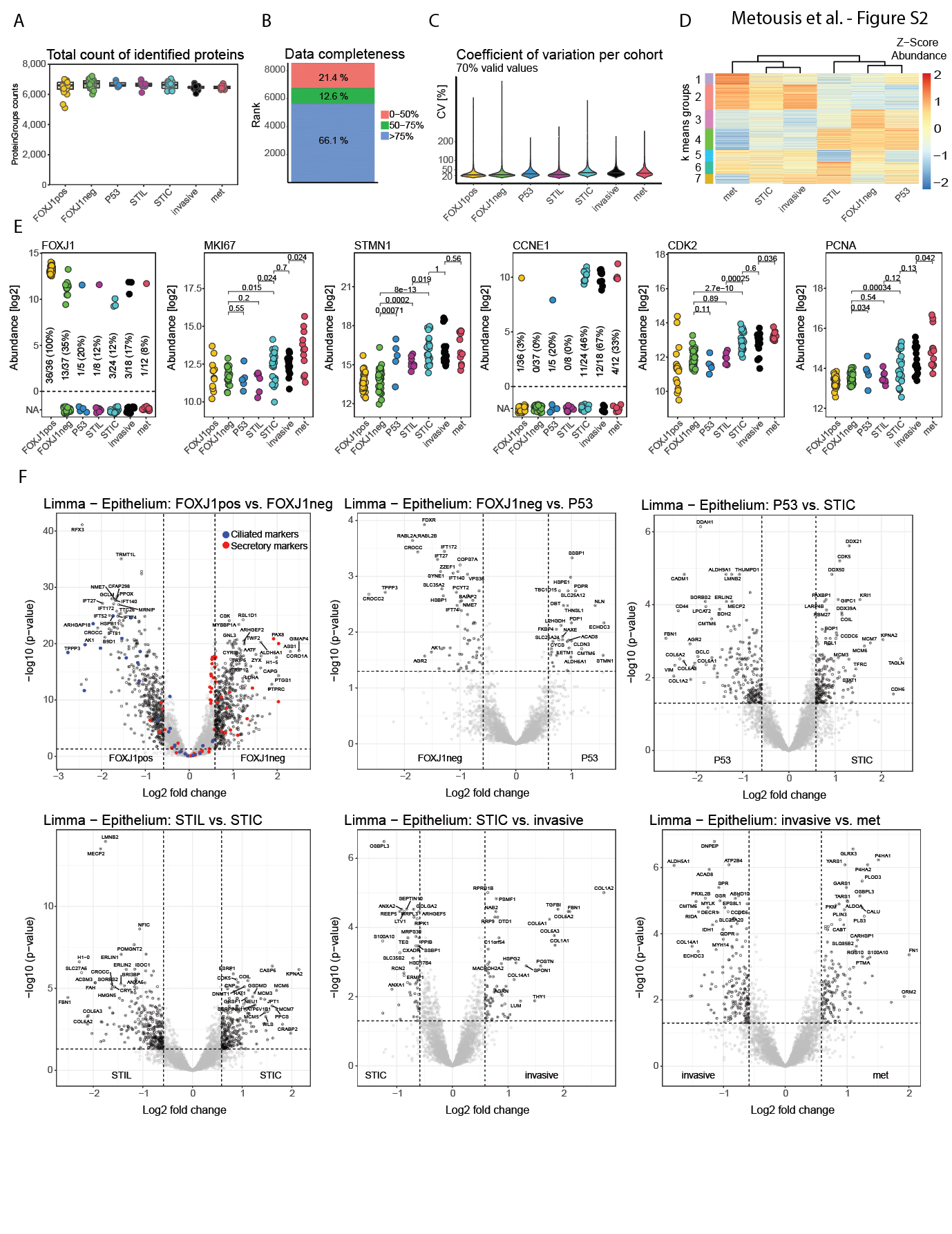
 **Figure S2. Quality control and differential protein expression in epithelial samples across the disease trajectory, related to Fig. 2**

(A) Total count of identified protein groups across different lesion types. Box plots show median, quartiles, and range; individual data points represent individual samples. FOXJ1pos, FOXJ1-positive cells. FOXJ1neg, FOXJ1-negative cells. P53, p53 signature. met, metastasis.

(B) Data completeness showing the percentage distribution of proteins with different coverage across all samples. The percentage of proteins identified in >75% (blue), 50-75% (green), and <50% (red) of all samples, respectively.

(C) Coefficient of variation (CV) for proteins across different lesion cohorts. CV distribution for proteins with 70-100% valid values. Violin plots display the density of CV values.

(D) K-means clustering analysis of epithelial proteins, excluding FOXJ1pos. Heatmap of normalized protein abundance (z-score) across different cell states. Scale indicates relative protein expression levels.

(E) Proteomics abundance of FOXJ1, Ki-67, STMN1, CCNE1, CDK2 and PCNA.

(F) Volcano plots showing differentially expressed proteins between sequential stages of disease progression (Table S2B). Gray dots represent proteins with non-significant changes; colored dots represent significantly differentially expressed proteins (red: secretory markers, blue: ciliated markers; FDR < 0.05, |fold change| >= 1.5). Key proteins are labeled.

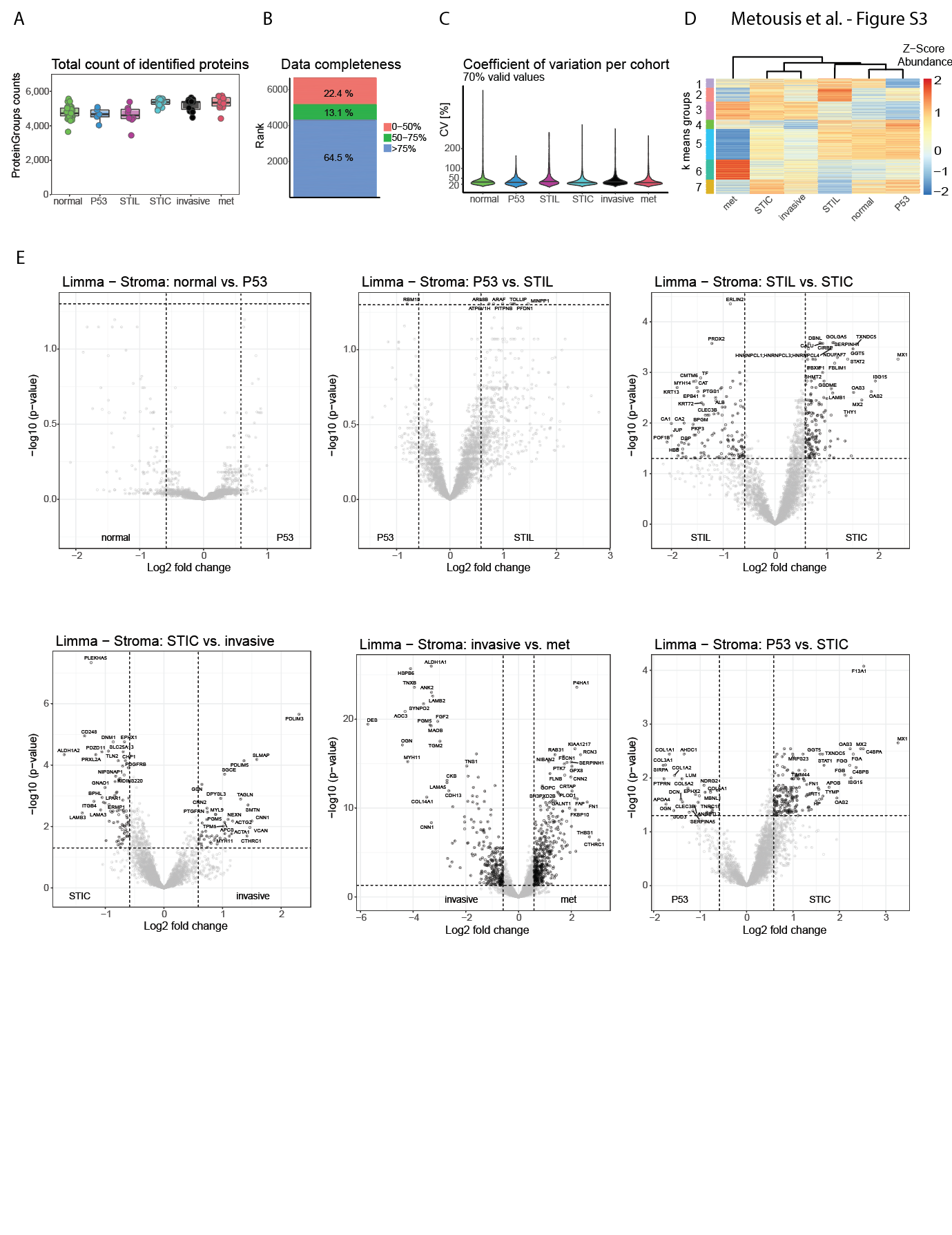

**Figure S3. Stromal proteome analysis across disease progression, related to Fig. 3**

(A) Total count of identified stromal protein groups across different lesion types. Box plots show median, quartiles, and range; individual data points represent individual samples. FOXJ1pos, FOXJ1-positive cells. FOXJ1neg, FOXJ1-negative cells. P53, p53 signature. met, metastasis.

(B) Data completeness showing the percentage distribution of proteins with different coverage across all samples. The percentage of proteins identified in >75% (blue), 50-75% (green), and <50% (red) of all samples, respectively.

(C) Coefficient of variation (CV) for proteins across different lesion cohorts. CV distribution for proteins with 70-100% valid values. Violin plots display the density of CV values.

(D) K-means clustering analysis of stromal proteins in seven clusters. Heatmap of normalized protein abundance (z-score) across different cell states. Scale indicates relative protein expression levels.

(E) Volcano plots showing differentially expressed proteins in the stroma below the epithelium over the preneoplastic progression series (Table S3B). Gray dots represent proteins with non-significant changes; black dots represent significantly differentially expressed proteins (FDR < 0.05, |fold change| > 1.5). Black dots represent significantly differentially expressed proteins (FDR < 0.05, |fold change| >= 1.5). Key proteins are labeled.

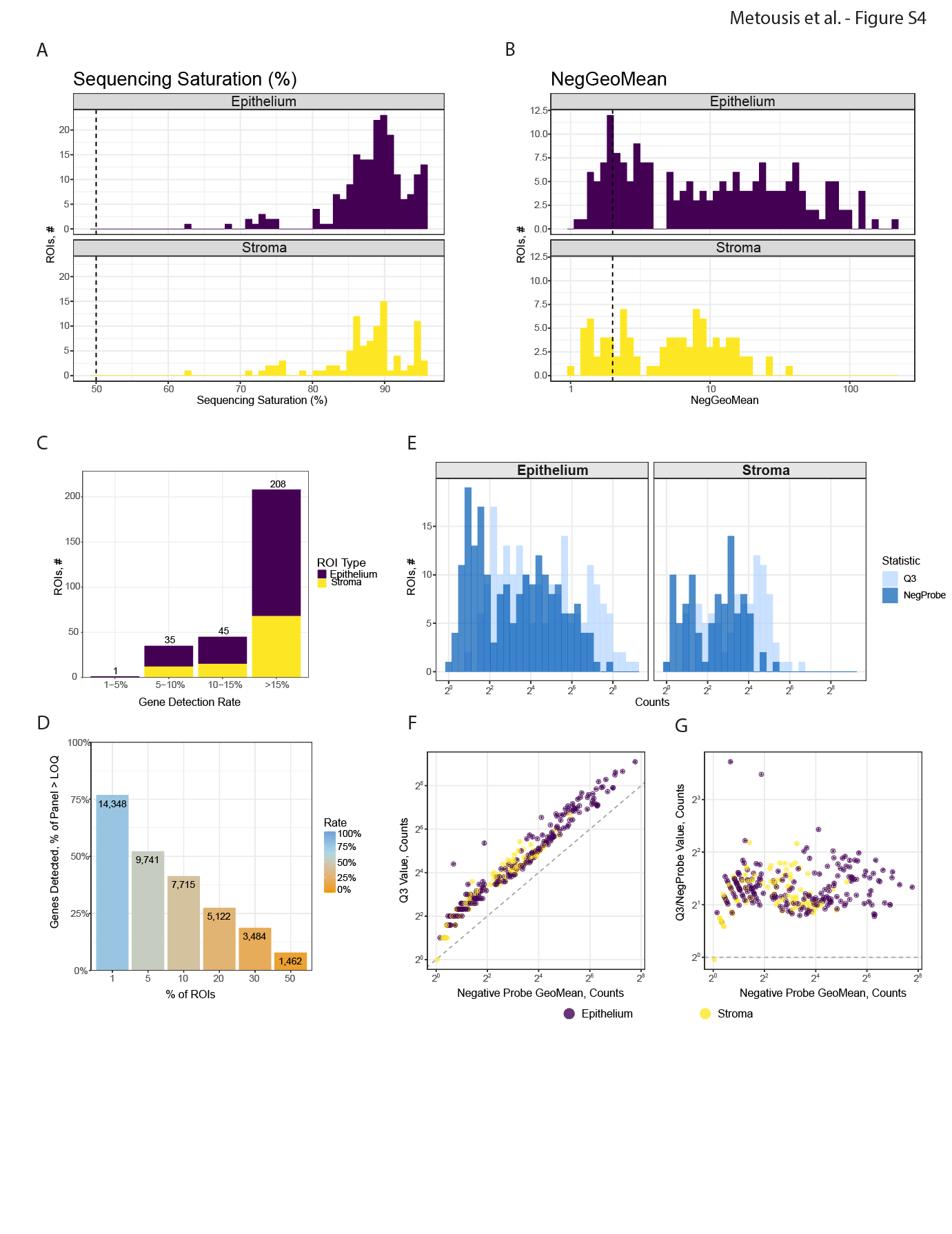
 **Figure S4. Quality control and normalization of spatial transcriptomics data, related to Fig. 4**

(A) Distribution of sequencing saturation rates for epithelial (purple) and stromal (yellow) samples. The dashed line indicates the 50% quality threshold used for sample filtering. ROI, region of interest.

(B) Distribution of negative geometric mean values (NegGeoMean) from Nanostring GeoMx transcriptomics quality control metrics for epithelial and stromal samples. Dashed line indicates the quality threshold applied during sample filtration.

(C) Sample distribution across different gene detection rate thresholds. Stacked bar chart shows the number of epithelial and stromal samples within each gene detection rate category. A total of 208 high-quality samples achieved >15% gene detection rate. ROIs < 5% gene detection rate were excluded from the analysis.

(D) Relationship between gene detection and percentage of ROIs. Bar chart shows the number of genes detected across different percentages of ROIs. Colors indicate detection rate from 100% (blue) to 0% (orange).

(E) Distribution of raw (light blue) and Q3-normalized (dark blue) counts, demonstrating effective normalization of the expression data.

(F) Correlation between negative probe geometric mean counts and Quartile 3 (Q3) values across epithelial and stromal samples, indicating proper technical correction during normalization.

(G) Relationship between negative probe geometric mean (GeoMean) counts and Q3/negative prove value counts, showing variable technical noise across samples addressed during normalization.

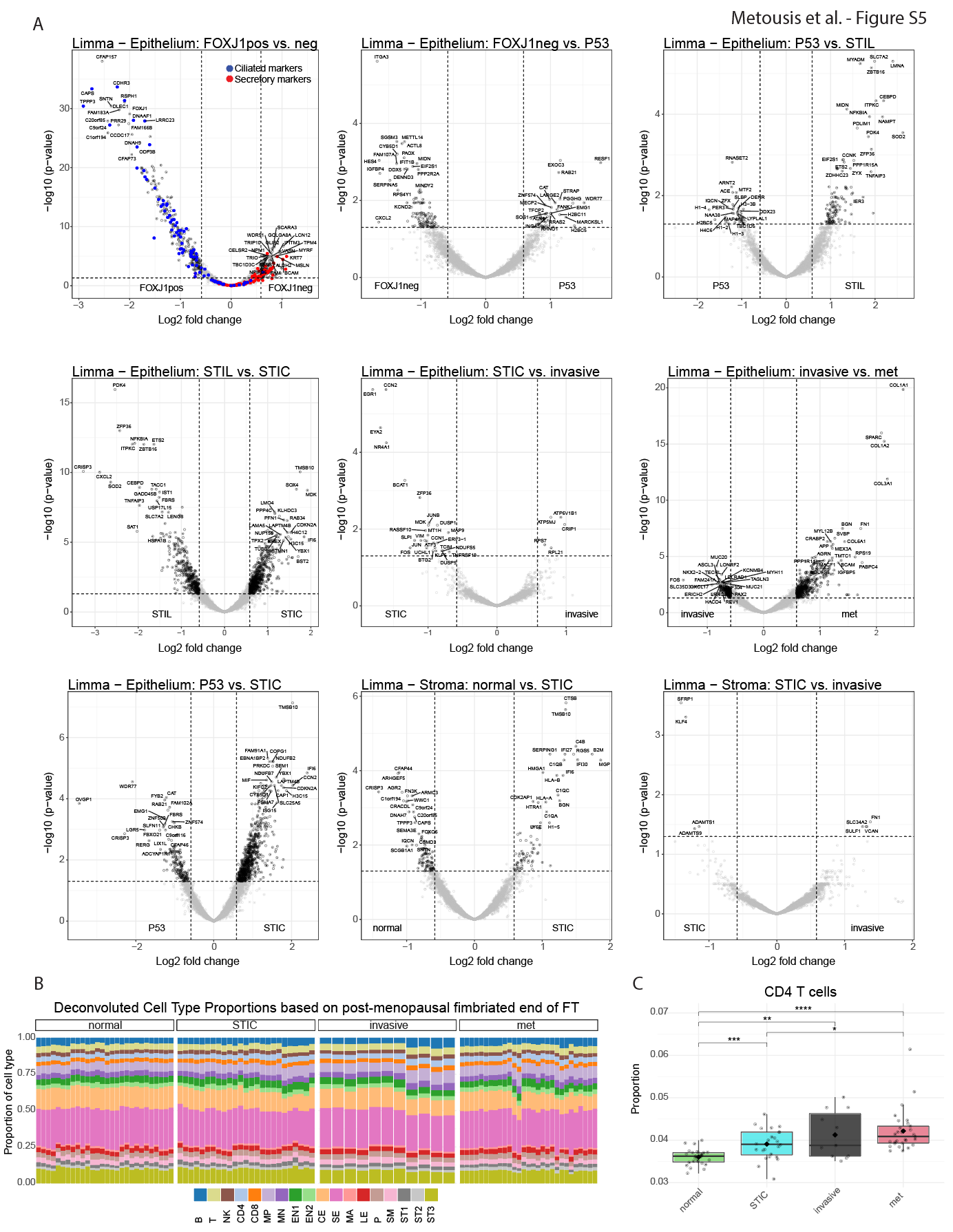
 **Figure S5. Epithelial and stromal transcriptome analysis across disease progression, related to Fig. 4**

(A) Volcano plots showing differentially expressed transcripts between sequential stages of disease progression (Table S4B, D). Gray dots represent proteins with non-significant changes; colored dots represent significantly differentially expressed proteins (red: secretory markers, blue: ciliated markers; FDR < 0.05, |fold change| >= 1.5). Key proteins are labeled. FOXJ1pos, FOXJ1-positive. FOXJ1neg, FOXJ1-negative. P53, p53 signature. met, metastasis.

(B) Stacked bar plot showing deconvoluted cell type proportions in fallopian tube (FT) stroma samples stratified by disease status using SpatialDecon^2^ (Table S4F). Due to limited sample numbers, the single p53 signature sample was grouped with normal samples, and the single STIL sample was grouped with STIC samples for visualization. Each vertical bar represents an individual sample, with colors indicating the relative proportions of different cell types identified through deconvolution analysis of Nanostring GeoMx transcriptomics data. Cell types are abbreviated as follows: B, B cells; T, T cells; NK, natural killer cells; CD4, CD4+ T cells; CD8, CD8+ T cells; MP, macrophages; MN, monocytes; EN1-2, endothelial cells subtypes 1-2; CE, ciliated epithelial cells; SE, secretory epithelial cells; LE, lymphatic endothelial cells; P, Pericytes; SM, smooth muscle cells; ST1-3, stromal cell subtypes that express stromelysin 1-3.

(C) Box plot showing the proportion of CD4+ T cells across disease states (Table S4G). Each dot represents an individual sample. Statistical significance was determined using Kruskal-Wallis test. **p < 0.01, ***p < 0.001, ****p < 0.0001. Box plots show median (center line), first and third quartiles (box limits), and 1.5× interquartile range (whiskers). Diamond shows mean.

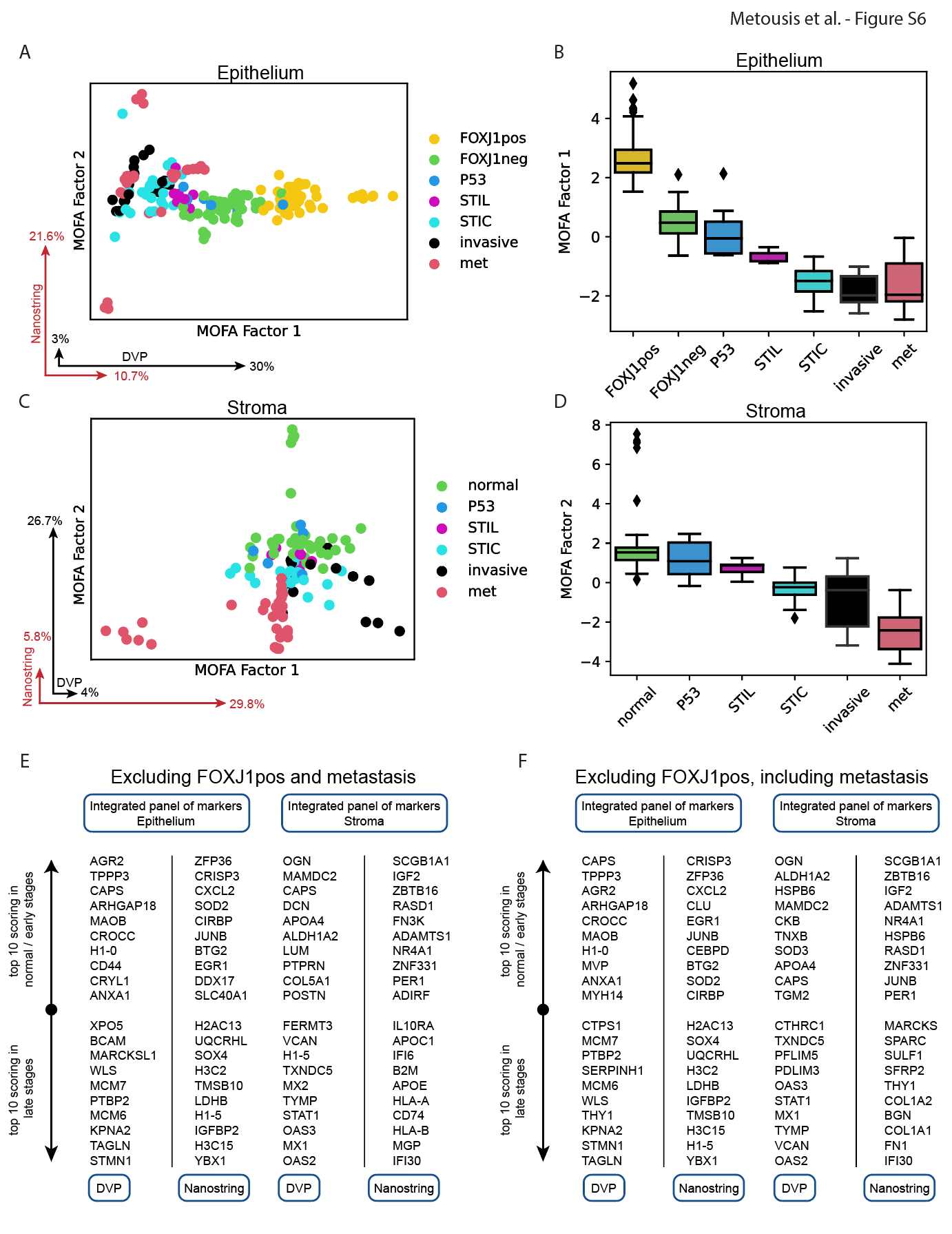
 **Figure S6. MOFA+ integration of spatial proteomics and transcriptomics data from secretory cells, related to Fig. 5**

(A-D). MOFA+ analysis, including metastasis (met) and FOXJ1-positive (FOXJ1pos). The first two MOFA factors for epithelial (A) and stromal (C) samples with points representing samples; colored by cell type/lesion state. Black and red vectors show the percentage of variability of deep visual proteomics (DVP) and Nanostring GeoMx transcriptomics (Nanostring) datasets captured by each MOFA factor. Box plots of MOFA factor 1 values across epithelial (B) and MOFA factor 2 values across stromal (D) cell states. FOXJ1pos, FOXJ1-positive. FOXJ1neg, FOXJ1-negative. P53, p53 signature. met, metastasis.

(E-F) Integrated panel of top molecular markers identified through combined DVP and Nanostring analysis excluding FOXJ1-positive (FOXJ1pos) and metastasis (E, Table S5E-H) or excluding FOXJ1pos and including metastasis (F, Table S5A-D). Left panels show epithelial markers and right panels show stromal markers. Within each compartment, molecules are grouped by data source (DVP or Nanostring) and sorted by their loading scores on MOFA factor 1 for both epithelium and stroma.

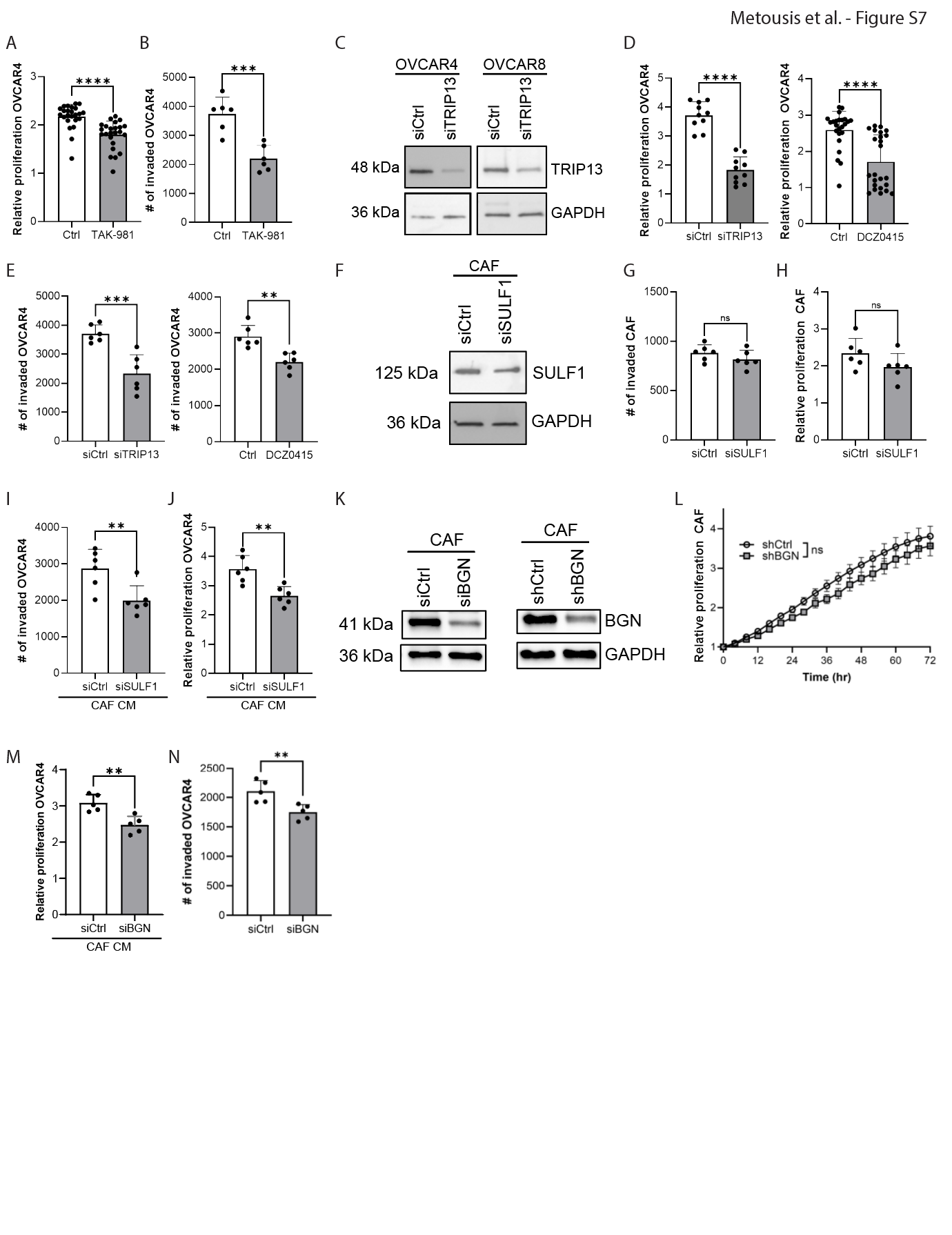
 **Figure S7. Functional validation of key molecular targets identified through spatial multi-omics, related to Fig. 6**

(A-B) OVCAR4 cells were treated with a SUMOylation inhibitor, TAK-981, and proliferation (A) or invasion (B) quantified.

(C) Immunoblot analysis of TRIP13. Lysates of OVCAR4 or OVCAR8 transfected with control siRNA (siCtrl), TRIP13 siRNA (siTRIP13) were collected. GAPDH serves as loading control.

(D-E) OVCAR4 cells were transfected with siCtrl or siTRIP (left) or treated with a TRIP13 inhibitor, DCZ0415 (right), and proliferation (D) or invasion (E) measured.

(F) Immunoblot analysis of sulfatase 1 (SULF1). Lysates of cancer-associated fibroblasts (CAFs) transfected with siCtrl or SULF1 siRNA (siSULF1) were collected. GAPDH serves as loading control.

(G-J) Cancer-associated fibroblasts (CAFs) were transfected with siCtrl or siSULF1 and proliferation (G), invasion (H), or conditioned-media (CAF CM) collected. OVCAR4 were treated with CAF CM and proliferation (I) or invasion (J) quantified.

(K) Immunoblot analysis of BGN. Lysates from CAFs transfected with siCtrl or BGN siRNA (siBGN; left), or CAFs stably expressing control shRNA (shRNA CAF) or BGN shRNA (shBGN CAF) were collected. GAPDH serves as loading control.

(L-N) CAFs were transfected with siCtrl or siBGN and CAF proliferation (L) or CAF conditioned-media (CAF CM) was collected. OVCAR4 cells were treated with the CAF CM, and proliferation (M) or invasion (N) quantified. Bar graphs show mean ± SD for three independent experiments: ns, not significant, ** p < 0.01, *** p < 0.001, **** p < 0.0001 by unpaired t-test.

### Supplementary tables

**Table S1. Demographics and cohort design, related to Fig. 1**

(A) Demographics of patient cohort.

(B) Study design for Deep Visual Proteomics.

(C) Study design for Nanostring.

(D) Mutations in homologous recombination repair genes.

**Table S2. Deep visual proteomics data on epithelial samples, related to Fig. 2**

(A) Normalized and log2 transformed deep visual proteomics dataset of epithelial samples.

(B) Differentially expressed proteins in epithelial compartments, related to Fig. 2H, S2F.

(C) K-means clusters of epithelial proteins, related to Fig. 2B.

(D) Gene set enrichment analysis (GSEA) on epithelial proteins from each K-means cluster.

(E) GSEA on differentially expressed epithelial proteins, related to Fig. 2C.

**Table S3. Deep visual proteomics data on stromal samples, related to Fig. 3**

(A) Normalized and log2 transformed deep visual proteomics dataset of stromal samples.

(B) Differentially expressed proteins in stromal compartments, related to Fig. 3H, S3E.

(C) K-means clusters of stromal proteins, related to Fig. 3B.

(D) Gene set enrichment analysis (GSEA) on stromal proteins from each K-means cluster.

(E) GSEA on differentially expressed stromal proteins, related to Fig. 3C.

**Table S4. Nanostring GeoMx transcriptomics data on epithelial and stromal samples, related to Fig. 4**

(A) Normalized and preprocessed Nanostring GeoMx transcriptomics dataset of epithelial and stromal samples.

(B) Differentially expressed transcripts in epithelial compartments, related to Fig. 4B, S5A.

(C) Gene set enrichment analysis (GSEA) on differentially expressed epithelial transcripts, related to Fig. 4C.

(D) Differentially expressed transcripts in stromal compartments, related to Fig. 4F, S5A.

(E) GSEA on differentially expressed stromal transcripts, related to Fig. 4G.

(F) Deconvoluted cell proportions per stromal transriptomic sample, related to Fig. S5B.

(G) Pairwise comparisons of deconvoluted cell proportions between sample types, related to Fig. 4J, S5C.

**Table S5. MOFA+ proteomics and transcriptomics data integration, related to Fig. 5**

(A) MOFA factor 1 values for proteins in the epithelium – excluding FOXJ1-positive samples, related to Fig. S6F.

(B) MOFA factor 1 values for transcripts in the epithelium – excluding FOXJ1-positive samples, related to Fig. S6F.

(C) MOFA factor 2 values for proteins in the stroma, related to Fig. S6F.

(D) MOFA factor 2 values for transcripts in the stroma, related to Fig. S6F.

(E) MOFA factor 1 values for proteins in the epithelium – excluding FOXJ1-positive and metastatic samples, related to Fig. S6E.

(F) MOFA factor 1 values for transcripts in the epithelium – excluding FOXJ1-positive and metastatic samples, related to Fig. S6E.

(G) MOFA factor 1 values for proteins in the stroma – excluding metastatic samples, related to Fig. S6E.

(H) MOFA factor 1 values for transcripts in the stroma – excluding metastatic samples, related to Fig. S6E.

(I) Gene set enrichment analysis (GSEA) on weighted proteins and transcripts in the epithelium – excluding FOXJ1-positive and metastatic samples, related to Fig. 5E.

(J) GSEA on weighted proteins and transcripts in the stroma – excluding metastatic samples, related to Fig. 5F.
